# Mental-health before and during the COVID-19 pandemic in adults with neurodevelopmental disorders

**DOI:** 10.1101/2022.05.09.22274714

**Authors:** Amy Shakeshaft, Rachel Blakey, Alex S.F. Kwong, Lucy Riglin, George Davey Smith, Evie Stergiakouli, Kate Tilling, Anita Thapar

## Abstract

The COVID-19 pandemic negatively impacted mental health globally. Individuals with neurodevelopmental disorders (NDDs), including autism spectrum disorder (ASD) and attention deficit hyperactivity disorder (ADHD), are at elevated risk of mental health difficulties. Therefore, we investigated the impact of the pandemic on anxiety, depression and mental wellbeing in adults with NDDs using longitudinal data from the Avon Longitudinal Study of Parents and Children study (n=3,058). Mental health data were collected pre-pandemic (age 21-25) and at three timepoints during the pandemic (ages 27-28) using the Short Mood and Feelings Questionnaire, Generalised Anxiety Disorder Assessment-7, and Warwick Edinburgh Mental Wellbeing Scale. ADHD and ASD were defined using validated cut-points of the Strengths and Difficulties Questionnaire and Autism Spectrum Quotient, self-reported at age 25. We used multi-level mixed-effects models to investigate changes in mental health in those with ADHD and ASD compared to those without. Prevalences of depression, anxiety and poor mental wellbeing were higher at all timepoints (pre-pandemic and during pandemic) in those with ADHD and ASD compared to those without. Anxiety increased to a greater extent in those with ADHD (β=0.8 [0.2,1.4], p=0.01) and ASD (β=1.2 [-0.1,2.5], p=0.07), while depression symptoms decreased, particularly in females with ASD (β=-3.1 [-4.6,-1.5], p=0.0001). On average, mental wellbeing decreased in all, but to a lesser extent in those with ADHD (β=1.3 [0.2,2.5], p=0.03) and females with ASD (β=3.0 [0.2,5.9], p=0.04). To conclude, anxiety disproportionately increased in adults with NDDs during the pandemic, however, the related lockdowns may have provided a protective environment for depressive symptoms in the same individuals.

## Introduction

The COVID-19 (SARS-CoV-2) pandemic has caused widespread disruption to lives worldwide. Virus suppression measures were introduced in many countries, with restrictions including orders to stay at home; school, workplace and business closures; and limited social mixing between households. The impact of the COVID-19 pandemic on mental health is now a widespread concern (Holmes et al., 2020), with a number of studies indicating substantial increases in mental ill-health (Robinson et al., 2022), particularly elevated anxiety, loneliness and psychological distress, and decreased life satisfaction and mental wellbeing throughout the pandemic (Kwong et al., 2020; Niedzwiedz et al., 2021; Patel et al., 2021; Santomauro et al., 2021). Despite the greater risk of severe physical illness from COVID-19 existing for older people (Pijls et al., 2021), population studies have highlighted the disproportionate effect of the pandemic on the mental health of younger people (Lewis et al., 2022; McGinty et al., 2020a; McGinty et al., 2020b; Niedzwiedz et al., 2021; Santomauro et al., 2021), likely influenced by job insecurity, financial pressures and education disruptions (Douglas et al., 2020; Gustafsson, 2020).

Some groups have been identified as especially vulnerable to poor mental health during the pandemic; for example, those from the poorest households (Pierre et al., 2021) and those with a history of previous mental health problems (Kwong et al., 2020; Pierce et al., 2021; Warne et al., 2021). Another potential risk group are individuals with neurodevelopmental disorders (NDDs), such as autism spectrum disorder (ASD) and attention deficit hyperactivity disorder (ADHD) who already have an elevated likelihood of mental health difficulties such as anxiety and depression (Faraone et al., 2021; Griffiths et al., 2019; Riglin et al., 2021b), as well as difficulties with work, education, romantic and social relationships (Farley et al., 2018; Riglin et al., *in press*; Thapar and Cooper, 2016) and thus may have fared especially poorly across pandemic restrictions.

Cross-sectional studies have suggested this may be the case. One mixed-method cross-sectional study indicated higher rates of depression and anxiety symptoms in adults with ASD compared to a non-ASD group during initial pandemic restrictions (Oomen et al., 2021) and suggested that loss of routine, loss of social contact, and the anticipation of life going back to normal were the most difficult and stressful changes for adults with ASD during restrictions. However, in the same study, some reported the reduced sensory and social overload during restrictions were a positive factor. Studies of children and adolescents during the pandemic show that those with NDDs or special educational needs had the highest levels of mental health difficulties (Creswell et al., 2021), with reports of the majority of children with ASD experiencing either a worsening of their pre-pandemic psychiatric diagnoses and/or the development of new psychiatric symptoms (Vasa et al., 2021). Whilst longitudinal studies in children and adolescents have indicated worse mental health outcomes in those with NDDs, the limited studies in adults with ADHD and ASD have lacked pre-pandemic baseline assessments (Lewis et al., 2022), did not use longitudinal study designs or did not have repeated measurements during the pandemic (Kwong et al., 2020).

In this study, we use a UK longitudinal birth cohort, ALSPAC, to investigate the mental health of individuals with NDDs, specifically ASD and ADHD, during the COVID-19 pandemic, comparing both to pre-pandemic levels, and to individuals without NDDs from the general population. Additionally, we investigate whether potentially influential circumstances during the pandemic, such as financial issues, family illness or COVID-19 infection, occurred as commonly in adults with NDDs as those without.

## Methods

### Sample

The Avon Longitudinal Study of Parents and Children (ALSPAC) study is an ongoing longitudinal study which recruited pregnant woman in the Avon region of South-West England, due to give birth between 1 April 1991 and 31 December 1992 (Boyd et al., 2013; Fraser et al., 2013; Northstone et al., 2019). The core sample consisted of 14,541 mothers and of these pregnancies, 13,988 children were alive at 1 year. Following the initial recruitment, an additional 913 children were recruited in three phases. Data were collected from families repeatedly until cohort members were age

25. Additional data were collected during the COVID-19 pandemic. Full details of the ALSPAC study and sample can be found in the Supplementary Material. For families with multiple births, we included the oldest sibling. We analysed data collected at ages 21, 23, 25 (pre-pandemic) and 27-28 (during pandemic).

### Measures and data collection

#### Pre-pandemic mental health

Anxiety symptoms were measured at age 21 using the self-rated Generalised Anxiety Disorder Assessment-7 (GAD-7) (Spitzer et al., 2006) (seven-items, range 0-21, with higher values indicating more generalised anxiety disorder symptoms). Mental wellbeing was measured at age 23 using the self-rated Warwick-Edinburgh Mental Wellbeing Scale (WEMWBS) (Tennant et al., 2007) (14-item questionnaire, range 14-70, with lower values indicating poorer mental wellbeing). Depressive symptoms were measured at age 25 using the self-rated Short Mood and Feelings Questionnaire (SMFQ) (Angold et al., 1995) (13-items, range 0-26, with higher values indicating more depressive symptoms). Total symptom scores were calculated for each scale to generate a continuous measure.

We also used recommended cut-points for these measures to estimate the number of individuals in the dataset with probable depression (SMFQ ≥12) (Eyre et al., 2021), generalised anxiety disorder (GAD-7 ≥10) (Spitzer et al., 2006), and poor mental wellbeing (WEMWBS ≤40) (Warwick Medical School., 2021).

#### Adult ADHD

To capture those with high ADHD symptoms, we used the self-rated 5-item Strengths and Difficulties Questionnaire (SDQ) (Goodman, 1997) ADHD subscale. This was completed by ALSPAC participants at age 25 years. This is designed to measure hyperactive and inattentive symptoms, ranges from 0-10, and has been validated against a DSM-5 diagnosis of ADHD (Riglin et al., 2021a), with a cut-point of ≥6 recommended to capture approximately 10% of the sample. Questionnaires with >2 items missing were excluded. Details of imputation of missing items is in Supplementary Material. Additional sensitivity analysis of SDQ ≥5 (recommended for high validity in correctly identifying those meeting diagnostic criteria in young adulthood) and ≥7 (recommended for use in younger ages respectively) cut-points (Riglin et al., 2021a): these are reported in Supplementary Material.

#### Adult ASD

To capture those with high ASD symptoms we used the self-rated Autism Spectrum Quotient (AQ). The AQ has been validated as a measure of clinical autism in adults, and ranges from 0-50, with a score of ≥32 indicating likely ASD (Baron-Cohen et al., 2001). This was completed by ALSPAC participants at age 25. Questionnaires with >10% (>5) missing items were not used. Details of imputation of missing items is in Supplementary Material. Additional sensitivity analysis of a broader cut-off point of AQ, suggested for screening of ASD (AQ ≥26 (Baron-Cohen et al., 2001)), is reported in Supplementary Material.

#### COVID-19 mental health

Questionnaires were sent to participants during (i) the first national lockdown in the UK from April to May 2020, (ii) when restrictions eased in the UK from May to July 2020 and (iii) during another national lockdown from December 2020 to January 2021. For specifics of UK restrictions see https://www.instituteforgovernment.org.uk/charts/uk-government-coronavirus-lockdowns. During these three questionnaire periods, all at ages 27-28 (referred to as COVID1, COVID2 and COVID3), depression was measured using the SMFQ, anxiety using the GAD-7 and mental wellbeing using the WEMWBS. Additionally, the presence or absence of potentially influential events since the start of the pandemic (March 2020), namely loss of job; financial difficulties; illness/injury of someone close to participant; or death of someone close to participant were recorded at the COVID3 timepoint.

#### COVID-19 infection

In addition to investigating mental health outcomes, we also used self-reported information recorded at the COVID3 timepoint on suspected or confirmed COVID-19 infections since the start of the pandemic, and the approximate date of infection, to investigate whether there were different distributions of infection in individuals with ASD/ADHD.

### Statistical Analysis Procedure

All analyses were conducted in R (R Core Team, 2021).

#### Mental health during COVID-19

The primary sample for analysis of mental health consisted of those with complete neurodevelopmental (AQ & SDQ), sex and COVID-19 event data (detailed above) and mental health outcome data from at least one timepoint (pre-pandemic, COVID1, COVID2 or COVID3). Previous studies have investigated the potential bias from attrition in the ALSPAC cohort with regards to mental health outcomes during the pandemic with complete case analysis showing near identical results to those using an imputed sample (Kwong et al., 2020), therefore motivating our choice of sample. The number of individuals and details of attrition are presented in Supplementary Figure 1. We initially describe cohort information, testing any differences between those with high vs. low ADHD/ASD symptoms using Chi-squared tests. We then present the prevalence of probable depression, generalised anxiety disorder and low mental wellbeing at pre-pandemic and COVID timepoints, stratified by those with ADHD/ASD. We also examined loss of job; financial difficulties; illness/injury of someone close to participant; or death of someone close to participant during the pandemic, and whether the frequency of these were different in those with ASD/ADHD compared to those without.

To investigate whether those with ADHD or ASD were differentially impacted by the pandemic, continuous mental health measures (SMFQ, GAD-7, WEMWBS) were modelled using multi-level mixed effects models using R2MLwiN (Zhang et al., 2016). These models account for correlations between repeated measures in the same individual and allow for different sample sizes at each timepoint, allowing maximisation of the possible sample without imputing missing outcome data. Three models were run for each mental health outcome: 1) with timepoints and ADHD/ASD symptoms included as fixed-effects; 2) sex-stratified models with timepoint and ADHD/ASD symptoms included and 3) models including timepoint, ADHD/ASD symptoms and sex with all possible interaction terms (2-way and 3-way) included. For all models, timepoints were included as dummy variables with the pre-pandemic timepoint as reference, ADHD or ASD symptoms were included as binary exposures based on cut-off points described above with low symptoms as reference, and sex was included with males as reference.

#### Sensitivity analysis

As there was an average of 6 years difference between pre-pandemic and pandemic GAD7 measurements, we tested for correlation between GAD7 at age 21 and an additional measure of anxiety, Screen for Adult Anxiety Related Disorders (SCAARED) (Angulo et al., 2017), at age 25, to ensure anxiety at age 21 was related with anxiety at age 25, just prior to the pandemic. We also repeated analysis of mental health outcomes using a variety of different cut-off points for ADHD and ASD, to ensure broader and narrower classifications showed the same pattern of results. These are reported in the Supplementary Material. We also describe changes in the mean scores for individual items in the GAD7 and SMFQ questionnaires in those with and without ADHD and ASD.

#### COVID-19 infection

For our second analysis of whether individuals with NDDs showed a different distribution of COVID-19 infection than those without, the sample included those with complete data on COVID-19 infection at COVID3 timepoint and complete ASD/ADHD exposure information. We first performed Chi-squared tests to determine whether the frequency of a suspected or confirmed COVID-19 infection by COVID3 timepoint was different in those with and without ASD or ADHD (as defined using the cut-offs described above). We then performed survival analysis using Kaplan-Meier

Estimation (Kaplan and Meier, 1958) and performed a log-rank test (Mantel, 1966) to test for differences in COVID-19 infection distribution between those with and without ADHD and ASD. The start date for survival analysis was set as 17^th^ November 2019 for all cases, estimated to be the first date of COVID-19 infection by Roberts et al. (2021). The date of COVID-19 infection was the event date and individuals were right-censored if they had not experienced an infection by the date of COVID3 survey completion.

## Results

### Description of sample before and during the COVID-19 pandemic

Data on anxiety, depression and mental wellbeing from at least one appropriate timepoint (pre-pandemic, COVID1, COVID2, COVID3) alongside complete neurodevelopmental data were available from 3,058 individuals (Supplementary Figure 1). Table 1 shows sample information. 386 individuals were classified with high ADHD symptoms at age 25 (13%; broadly defined ADHD) and 79 (3%) with high ASD symptoms (broadly defined ASD). Individuals with broadly defined ADHD were more likely to have experienced financial difficulties (OR = 1.34 [1.06,1.69], p=0.01) and the death of someone close (OR = 1.33 [1.02,1.72], p=0.03) during the pandemic (between March 2020 and March 2021).

**Table 1.** Description of total sample and those with ADHD and ASD with p-values from Chi-squared tests between ADHD (yes vs. no) and ASD (yes vs. no).

|  | Total sample | ADHD (SDQ ≥ 6) |  |  | ASD (ASQ ≥ 32) |  |  |
| --- | --- | --- | --- | --- | --- | --- | --- |
|  |  | Yes | No | P-value | Yes | No | P-value |
| **N** | 3,058 (100%) | 386 (13%) | 2672 (87%) | - | 79 (3%) | 2979 (97%) | - |
| **Sex** |  |  |  |  |  |  |  |
| Females (%) | 2,103 (69%) | 240 (62%) | 1,863 (70%) | **0.003** | 53 (67%) | 2,050 (69%) | 0.84 |
| **ADHD symptoms** |  |  |  |  |  |  |  |
| Mean SDQ score (SD) | 3.04 (2.10) | 6.82 (0.99) | 2.49 (1.60) | - | 4.82 (2.46) | 2.99 (2.07) | - |
| ADHD based on SDQ ≥ 6 (%) | 386 (13%) | 386 (100%) | 0 (0%) | - | 33 (42%) | 353 (12%) | **2.0x10<sup>-10</sup>** |
| **ASD symptoms** |  |  |  |  |  |  |  |
| Mean AQ score (SD) | 15.99 (7.19) | 20.73 (7.97) | 15.31 (6.80) | - | 35.61 (3.74) | 15.47 (6.50) | - |
| ASD based on AQ ≥ 32 (%) | 79 (3%) | 33 (9%) | 46 (2%) | **2.0x10<sup>-10</sup>** | 79 (100%) | 0 (0%) | - |
| **Events during pandemic** |  |  |  |  |  |  |  |
| Financial difficulties (%) | 764 (25%) | 116 (30%) | 648 (24%) | **0.01** | 23 (29%) | 741 (25%) | 0.39 |
| Lost job (%) | 239 (8%) | 37 (10%) | 202 (8%) | 0.17 | 6 (8%) | 233 (8%) | 0.94 |
| Death of someone close (%) | 533 (17%) | 82 (21%) | 451 (17%) | **0.03** | 10 (13%) | 523 (18%) | 0.26 |
| Illness/injury of someone close (%) | 514 (17%) | 63 (16%) | 451 (17%) | 0.78 | 10 (13%) | 504 (17%) | 0.32 |
| **Anxiety (GAD7)** |  |  |  |  |  |  |  |
| Pre-pandemic (%) | 255/1998 (13%) | 48/224 (21%) | 207/1774 (12%) | **3.7x10<sup>-5</sup>** | 12/47 (26%) | 243/1951 (12%) | **0.008** |
| COVID1 (%) | 491/2048 (24%) | 91/244 (37%) | 400/1804 (22%) | **2.1x10<sup>-7</sup>** | 22/53 (42%) | 469/1995 (24%) | **0.002** |
| COVID2 (%) | 437/1897 (23%) | 99/241 (41%) | 338/1656 (20%) | **1.1x10<sup>-12</sup>** | 25/54 (46%) | 412/1843 (22%) | **3.8x10<sup>-5</sup>** |
| COVID3 (%) | 820/3028 (27%) | 161/382 (42%) | 659/2646 (25%) | **1.4x10<sup>-12</sup>** | 37/79 (47%) | 783/2949 (27%) | **6.2x10<sup>-5</sup>** |
| **Depression (SMFQ)** |  |  |  |  |  |  |  |
| Pre-pandemic (%) | 615/2973 (21%) | 171/378 (45%) | 444/2595 (17%) | **1.8x10<sup>-36</sup>** | 47/77 (61%) | 568/2896 (20%) | **8.2x10<sup>-19</sup>** |
| COVID1 (%) | 299/2024 (15%) | 69/240 (29%) | 230/1784 (13%) | **8.0x10<sup>-11</sup>** | 21/52 (40%) | 278/1972 (14%) | **1.3x10<sup>-7</sup>** |
| COVID2 (%) | 340/1892 (18%) | 83/237 (35%) | 257/1655 (16%) | **2.6x10<sup>-13</sup>** | 30/56 (54%) | 310/1836 (17%) | **1.9x10<sup>-12</sup>** |
| COVID3 (%) | 558/2997 (19%) | 127/382 (33%) | 431/2615 (16%) | **3.8x10<sup>-15</sup>** | 33/78 (42%) | 525/2919 (18%) | **5.1x10<sup>-8</sup>** |
| **Poor mental wellbeing (WEMWBS)** |  |  |  |  |  |  |  |
| Pre-pandemic (%) | 413/2192 (19%) | 92/270 (34%) | 321/1922 (17%) | **8.2x10<sup>-12</sup>** | 25/59 (42%) | 388/2133 (18%) | **2.8x10<sup>-6</sup>** |
| COVID1 (%) | 628/2035 (31%) | 113/240 (47%) | 515/1795 (29%) | **6.9x10<sup>-9</sup>** | 31/51 (61%) | 597/1984 (30%) | **2.8x10<sup>-6</sup>** |
| COVID2 (%) | 602/1902 (32%) | 118/238 (50%) | 484/1664 (29%) | **2.0x10<sup>-10</sup>** | 33/55 (60%) | 569/1847 (31%) | **4.5x10<sup>-6</sup>** |
| COVID3 (%) | 928/3014 (31%) | 189/382 (49%) | 739/2632 (28%) | **2.5x10<sup>-17</sup>** | 46/77 (60%) | 882/2937 (30%) | **2.4x10<sup>-8</sup>** |

The prevalence of anxiety, depression and poor mental wellbeing based on validated cut-off thresholds at each timepoint, are presented in Table 1. The proportion of those with ASD and ADHD with anxiety, depression and poor mental wellbeing were consistently higher than those without at each timepoint. The prevalence of anxiety in those with ADHD increased from 21% pre-pandemic to 42% by COVID3, and from 26% to 47% across the same timeframe in those with ASD. Depression prevalence fell from 45% to 33% in those with ADHD, and 61% to 42% in those with ASD, while poor mental wellbeing increased from 34% to 49% in ADHD and 42% to 60% in ASD.

### Changes in mental health in ADHD and ASD during the COVID-19 pandemic

#### ADHD

Anxiety symptoms were consistently higher in those with ADHD compared to those without at all timepoints (Figure 2). Anxiety was higher in those with and without ADHD at all COVID timepoints compared to pre-pandemic levels, but the increase in anxiety from the pre-pandemic level was elevated in those with ADHD at COVID2 (COVID2*ADHD β = 0.8 [0.1, 1.5], p=0.03) and COVID3 (COVID3*ADHD β= 0.8 [0.2, 1.4], p=0.01) timepoints (Figure 3, Supplementary Table 1).

**Figure 1.**
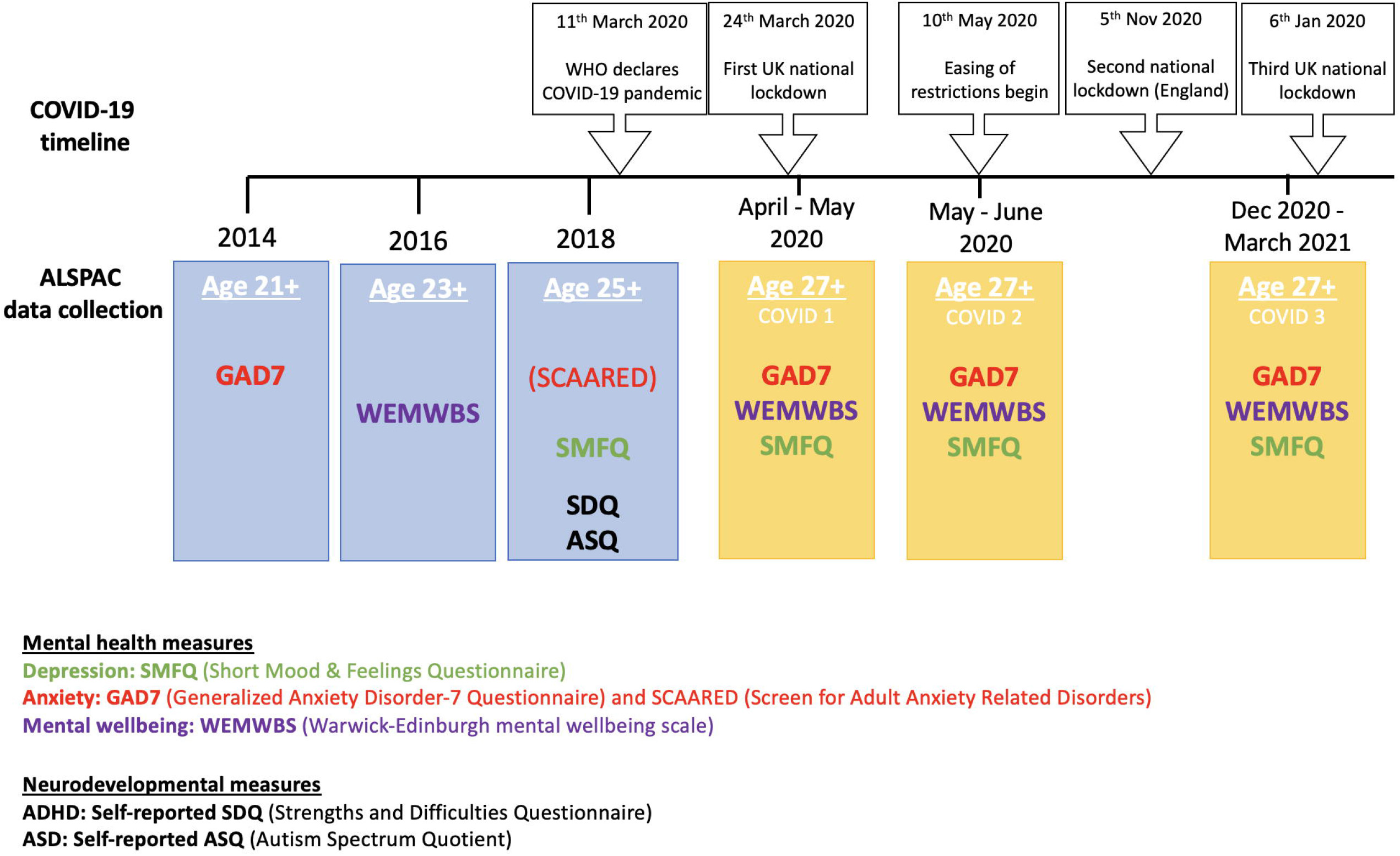
Timeline of ALSPAC data collection relating to mental health and major COVID-19 events in the UK. World Health Organisation (WHO).

**Figure 2.**
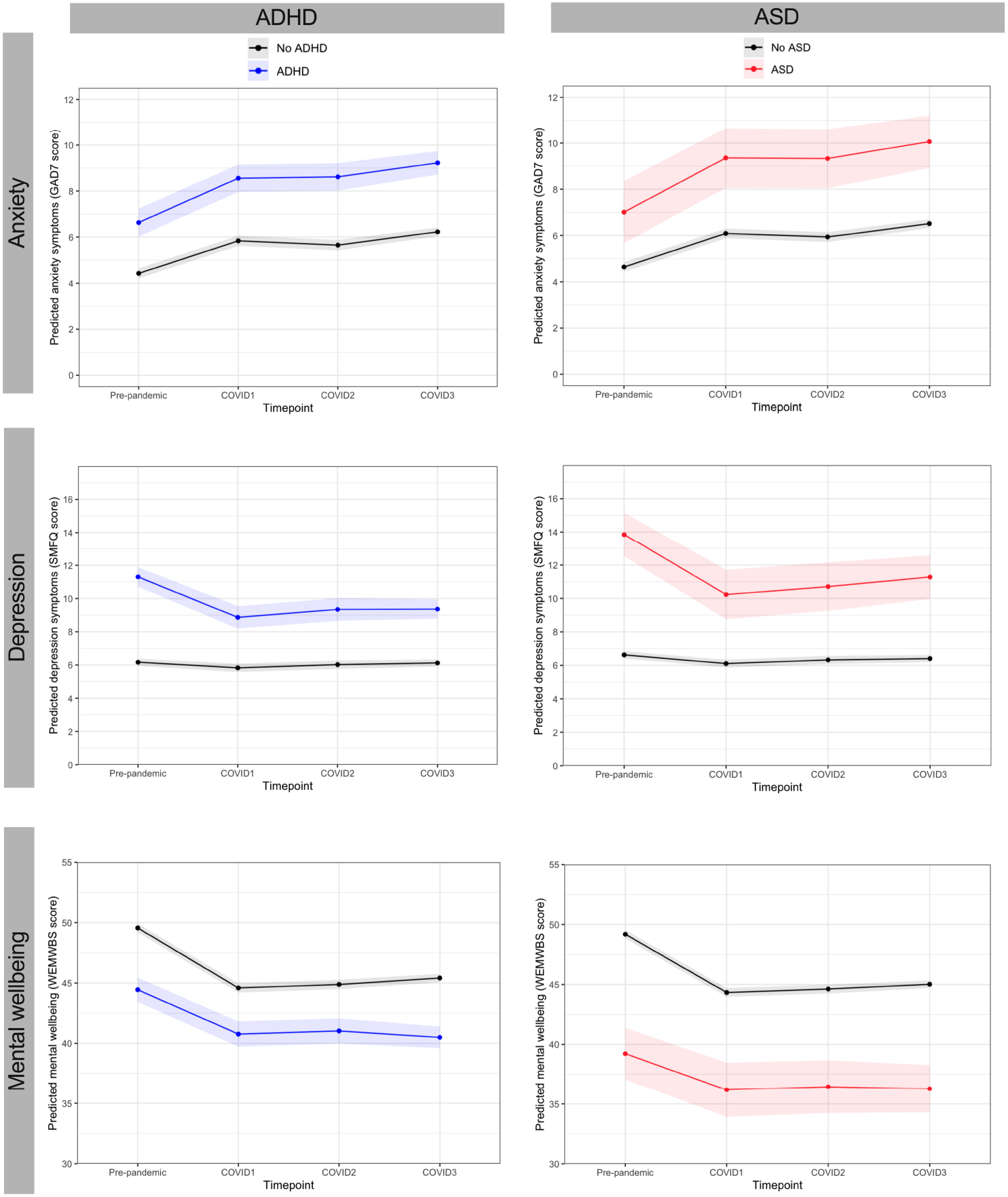
Trajectories of anxiety, depression and mental wellbeing during COVID-19 pandemic in those with and without ADHD and ASD. Shaded regions represent 95% confidence intervals.

**Figure 3.**
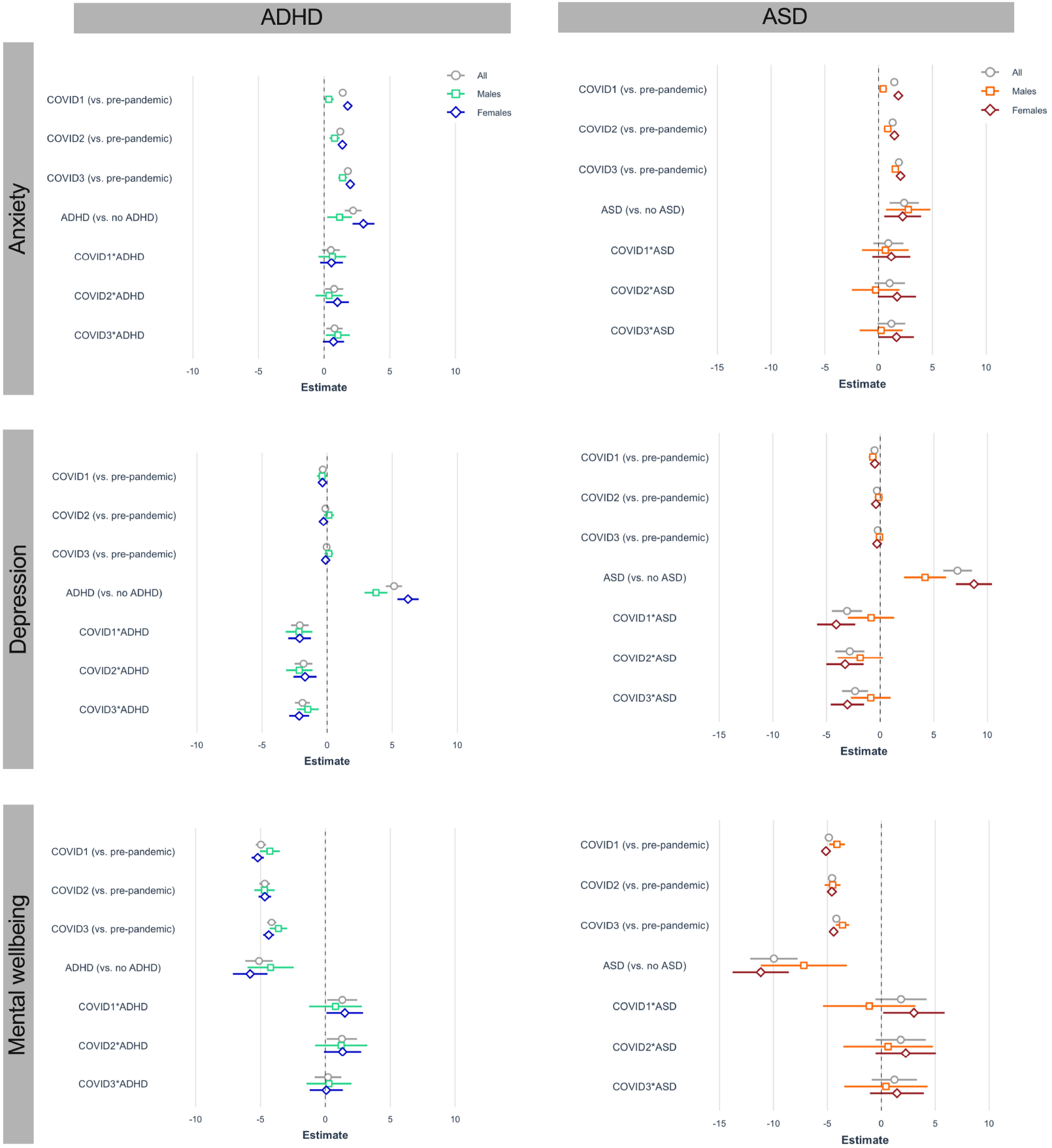
Results from multi-level models of mental health outcomes during the COVID-19 pandemic. Unstandardised β coefficient estimates with 95% confidence intervals from total and sex-stratified models are shown. P-values for estimates are presented in the Supplementary Material.

Depression symptoms were consistently higher in those with ADHD compared to those without (Figure 2). During the pandemic, symptoms were lower than pre-pandemic levels in those with ADHD, whereas in those without ADHD, depressive symptoms stayed relatively stable. The decrease in depression between pre-pandemic and all 3 COVID timepoints was significantly greater in those with ADHD than without (COVID1*ADHD β= -2.1 [-2.8,-1.4], p=1.5×10^−9^; COVID2*ADHD β=-1.8 [-2.5,-1.1], p=2.1×10^−7^; COVID3*ADHD β=-1.9 [-2.5,1.3], p=1.5×10^−10^) (Figure 3, Supplementary Table 3).

Mental wellbeing was consistently lower in those with ADHD compared to those without at each timepoint (Figure 2), and was lower in those with and without ADHD at all COVID timepoints compared to pre-pandemic. The decrease in mental wellbeing from pre-pandemic levels was to a lesser extent in those with ADHD at COVID1 (COVID1*ADHD β=1.3 [0.2, 2.5], p=0.03) and COVID2 (COVID2*ADHD β=1.3 [0.1, 2.4], p=0.03) timepoints (Figure 3, Supplementary Table 5).

Comparison of mean scores for each item of GAD7 and SMFQ in those with and without ADHD are presented in Supplementary Figure 2 and 3. All GAD7 items indicated worse anxiety symptoms during the pandemic compared to pre-pandemic in those with and without ADHD. For SMFQ, most items (8/13) indicated better (less) depressive symptoms during the pandemic. The item ‘felt no enjoyment’ indicated worse symptoms during the pandemic in both those with and without ADHD. Other items, such as ‘feeling miserable’, ‘feeling lonely’ and ‘finding it hard to think’ slightly improved on average in those with ADHD during the pandemic but got slightly worse in those without ADHD.

#### ASD

The pattern of mental health trajectories among people with ASD and ADHD were similar. Anxiety symptoms were higher in those with ASD compared to those without at all timepoints, and higher in those with and without ASD during the pandemic compared to pre-pandemic levels (Figure 2), showing some evidence of an elevated change compared to pre-pandemic levels in ASD (COVID3*ASD β=1.2 [-0.1,2.5], p=0.07) (Figure 3, Supplementary Table 2).

Depression symptoms were consistently higher in those with ASD at each timepoint. During the pandemic, depression symptoms were lower than pre-pandemic levels in those with ASD, whereas in those without ASD, symptoms stayed relatively stable (Figure 2). This decrease in depression between pre-pandemic and COVID timepoints was greater in those with ASD than without (COVID1*ASD β=-3.1 [-4.5,8.6], p=1.6×10^−5^; COVID2*ASD β=-2.8 [-4.2,1.5], p=4.9×10^−5^; COVID3*ASD β=-

2.3 [-3.6,-1.1], p=0.0002) (Figure 3, Supplementary Table 4).

Mental wellbeing was consistently lower in those with ASD compared to those without at each timepoint, and was lower on average in both groups during the pandemic compared to pre-pandemic (Figure 2). There was some evidence that mental wellbeing decreased to a lesser extent from pre-pandemic levels in those with ASD compared to those without (COVID1*ASD β=1.82 [-0.6,4.2], p=0.13) (Figure 3, Supplementary Table 6).

Comparison of mean scores for each item of GAD7 and SMFQ in those with and without ASD are presented in Supplementary Figure 2 and 3. All GAD7 items indicated worse anxiety symptoms during the pandemic compared to pre-pandemic in those with and without ASD. For SMFQ, most items (7/13) indicated better (less) depressive symptoms during the pandemic. The item ‘felt no enjoyment’ indicated worse symptoms during the pandemic in both those with and without ASD. Other items, such as ‘feeling miserable’, ‘feeling restless’, ‘feeling lonely’ and ‘finding it hard to think’ slightly improved on average in those with ASD during the pandemic but got slightly worse in those without ASD.

#### Sex differences in mental health

Figure 3 shows β coefficients from models stratified by sex as well as in the whole cohort, indicating the presence of sex differences in changes in anxiety, depression and mental wellbeing in ASD. The increase in anxiety at COVID2 and 3 is more pronounced in females with ASD (COVID2*ASD β=1.7 [-0.6,3.9], p=0.06; COVID3*ASD β=1.7 [0.03,3.3], p=0.05) compared to males with ASD (COVID2*ASD β=-0.3 [-2.5,1.9], p=0.8; COVID3*ASD β=0.2 [-1.8,2.2], p=0.8). Depression decreases to a greater extent (greater improvement compared to pre-pandemic levels) and mental wellbeing decreases to a lesser extent (stays closer to pre-pandemic levels) in females with ASD at all three COVID timepoints. Inclusion of a 3-way interaction term in the model between COVID timepoints, ASD and sex (Supplementary Table 4) showed an effect modification of sex on the interaction between depression at COVID 1 and ASD, with females with ASD showing a greater decline in depressive symptoms compared to other strata (COVID1*ASD*Sex β=-3.3 [-6.3, -0.3], p=0.03).

Sex-stratified mental health trajectories in those with and without ADHD and ASD are shown in Supplementary Figure 4. Females with ADHD/ASD had consistently the poorest mental health out of all strata at each timepoint investigated, and males without ADHD/ASD consistently had the best.

#### Sensitivity analysis

There was a large correlation between GAD7, at age 21, and SCAARED, at age 25 (Spearman’s r=0.54, p=2.2×10^−16^). The effects of ADHD and ASD on changes in mental health remained very similar when using broader and narrower cut-off points for the SDQ and AQ (Supplementary Figures 5 and 6).

### COVID-19 infection

Survival analysis did not show strong evidence of differences in the distribution of infections in those with ADHD (χ^2^(1)=1.1, p=0.29) or ASD (χ^2^(1)=1.7, p=0.19) compared to those without (Supplementary Material).

## Discussion

In this study we investigate the impact of the COVID-19 pandemic on the mental health of young adults with NDDs. We show, on average, a substantial increase in anxiety during the pandemic among those with ADHD and ASD compared to those without. In contrast, mean depression symptoms improved in those with ADHD and in females with ASD, whilst remaining unchanged in those without. Mental wellbeing declined on average among all individuals, but this was to a lesser extent in those with ADHD and females with ASD.

It is commonly reported that individuals with NDDs such as ADHD and ASD are at an elevated likelihood to experience mental health difficulties, especially anxiety and depression, during the lifespan (Faraone et al., 2021; Griffiths et al., 2019; Riglin et al., 2021b). Results from this study indicate that the prevalence of anxiety, depression and poor mental wellbeing remain well above the population prevalence both before and during the pandemic, with 37-47% of individuals with ADHD and ASD being classified as having a possible generalised anxiety disorder during the pandemic compared to between 20-27% of those without. The global increase in anxiety during the pandemic has attracted considerable concern, and here we show that this increase in anxiety was particularly marked for young adults with ASD and ADHD in the UK. Factors likely contributing to this increased anxiety include job instability and financial difficulties which were reported in around one-third of participants with ASD and ADHD compared to one-quarter of those without. Results from our survival analysis indicate that differences in mental health changes during the pandemic in those with ADHD and ASD were unlikely due to differing COVID-19 infections rates between groups.

In the case of depression, previous studies show little change in depressive symptoms during the pandemic in the ALSPAC cohort (Kwong et al., 2020). This contrasts to evidence from a recent meta-analysis of longitudinal studies which showed an overall increase in depression and mood disorder symptoms during the pandemic (Robinson et al., 2022). However, data from our study indicates that, while the prevalence of depression remains higher in those with NDDs during the pandemic (up to 54% of those with ASD and 35% of those with ADHD), depressive symptoms actually improved compared to pre-pandemic levels. For those with ADHD and females with ASD, depressive symptoms are the lowest during the first COVID timepoint, when the UK was in strictest national lockdown, with schools and many workplaces closed and limitations in place on socialising and leaving the house. Similarly, a longitudinal study in Western Australia showed that children and adolescents with ADHD reported decreases in depression and increases in positive wellbeing between pre-COVID and school closures due to the pandemic (Houghton et al., 2022). Qualitative studies in adults and children with ADHD have reported some positives of lockdown, including the flexibility of managing schedules, and less exposure to negative feedback (Ando et al., 2021; Bobo et al., 2020). Additionally, some individuals with ASD reported that the relief of day-to-day social demands was appreciated during the early lockdown, with less demands to camouflage or mask behaviour (Bundy et al., 2022). This potentially protective environment of lockdown may be particularly prevalent for females with ASD, for whom we saw the greatest decrease in depressive symptoms. Previous evidence indicates that females with ASD commonly mask or camouflage behaviours in social situations in order to ‘fit in’ (Hull et al., 2019; Hull et al., 2017) and camouflaging has been linked to greater symptoms of depression and anxiety in autistic adults (Hull et al., 2021). Therefore, it may be possible that the lack of expectation to socialise during the first COVID lockdown eased the pressure for females with ASD to camouflage autistic characteristics.

The opposite direction between changes in anxiety vs. depression symptoms in individuals with ADHD and ASD was somewhat unexpected given the strong correlation between these two aspects of mental health and their known comorbidity (Johansson et al., 2013). However, evidence suggests that anxiety often precedes and can predict depression up to a decade later (Davies et al., 2016; Jacobson and Newman, 2014), and that anxiety may change more rapidly than depression (in the case of positive changes after treatment) (Lewis et al., 2019). Therefore, it may be too soon to predict the effect of the pandemic on long-term mental health outcomes.

Whilst COVID-19 restrictions have mostly eased in the UK and many other countries at the time of writing, the results presented in this study, in addition to other studies, highlight the widespread effect the pandemic has had on mental health, particularly anxiety. This study indicates a disproportionate effect on anxiety in adults with NDDs which should be considered with regards to provision of support, both in the case of further restrictions and for management of mental health recovery in those most impacted (Vadivel et al., 2021).

This study has several important strengths. The longitudinal study design means robust pre-pandemic measures of mental health were present for comparison, preventing any chance of recall bias which may be present in cross-sectional or retrospective data collection. Also, multiple waves of data collection enabled us to capture mental health changes at different stages of national lockdowns. Further, by using multi-level models we were able to maximise the sample to include all available data.

The results of this study should be viewed considering methodological limitations. Like many longitudinal population-based studies, ALSPAC suffers from non-random attrition, with those at elevated risk of psychopathology, including those with high genetic risk for ADHD and depression, being more likely to drop-out (Taylor et al., 2018). Further, in this study we focused on ADHD and ASD symptoms in a population cohort, and although evidence suggests that both traits are continuously distributed (Larsson et al., 2012; Rutter and Pine, 2015; Thapar and Cooper, 2016), our findings may have limited generalisability to clinical samples of ADHD and ASD which have strict diagnostic criteria. The limited sample sizes for NDD groups, especially ASD (n=79, 3%), is also a limitation, and contributes to wider confidence intervals for mental health estimates. The different timings of pre-pandemic measures of depression (age 25), mental wellbeing (age 23) and anxiety (age 21) are not ideal for assessing change from baseline, nor the 6-year gap between pre-pandemic and pandemic anxiety measures. However, we saw high correlation between anxiety scores at age 21 and at age 25 using SCAARED, indicating that this pre-pandemic measure is still acceptable for comparison. Further, we note that we cannot assess whether changes in mental health observed are a direct result of the pandemic rather than a result of the natural trajectory of mental health, given the lack of comparison data on mental health trajectories in a non-pandemic exposed young adulthood population from this generation.

## Conclusion

Prior to the COVID-19 pandemic individuals with NDDs were at heightened risk of poor mental health, but this study indicates this risk has substantially grown during the pandemic and associated lockdown measures. Risk factors for poor mental health including experiencing financial difficulties and death of someone close were also greater in those with NDDs during this period. Anxiety increased to a greater degree in young adults with ASD and ADHD compared to those without, and mental wellbeing also decreased. The decline in depressive symptoms observed in individuals with ADHD and females with ASD raises issues surrounding the contribution of daily pressures such as camouflaging behaviours and maintaining strict routines to the development of poor mental health in adults with NDDs.

## Supporting information

STROBE checklist

## Data Availability

Any researcher can apply to use ALSPAC data, including the variables under investigation in this study. Access information is provided here: http://www.bristol.ac.uk/alspac/researchers/access/

## Author Contributions

**Amy Shakeshaft**: Methodology, Formal analysis, Investigation, Visualization, Writing - original draft, Writing - Review & editing; **Alex SF Kwong**: Methodology, Investigation, Writing - review & editing; **Rachel Blakey**: Investigation, Methodology, Writing - review & editing; **Lucy Riglin**: Methodology, Conceptualization, Supervision, Writing - review & editing; **George Davey Smith**: Funding acquisition, Writing - review & editing; **Kate Tilling**: Conceptualization, Methodology, Funding acquisition, Project administration, Supervision, Writing - review & editing; **Evie Stergiakouli**: Conceptualization, Funding acquisition, Project administration, Supervision, Writing - review & editing; **Anita Thapar**: Conceptualization, Funding acquisition, Project administration, Supervision, Writing - review & editing

## Acknowledgements

We are extremely grateful to all the families who took part in this study, the midwives for their help in recruiting them, and the whole ALSPAC team, which includes interviewers, computer and laboratory technicians, clerical workers, research scientists, volunteers, managers, receptionists and nurses.

## References

Ando, M., Takeda, T., Kumagai, K., 2021. A Qualitative Study of Impacts of the COVID-19 Pandemic on Lives in Adults with Attention Deficit Hyperactive Disorder in Japan. Int J Environ Res Public Health 18(4).

Angold, A., Costello, E.J., Messer, S.C., Pickles, A., 1995. Development of a short questionnaire for use in epidemiological studies of depression in children and adolescents. International journal of methods in psychiatric research.

Angulo, M., Rooks, B.T., Gill, M., Goldstein, T., Sakolsky, D., Goldstein, B., Monk, K., Hickey, M.B., Diler, R.S., Hafeman, D., Merranko, J., Axelson, D., Birmaher, B., 2017. Psychometrics of the screen for adult anxiety related disorders (SCAARED)-A new scale for the assessment of DSM-5 anxiety disorders. Psychiatry Res 253, 84–90.

Baron-Cohen, S., Wheelwright, S., Skinner, R., Martin, J., Clubley, E., 2001. The autism-spectrum quotient (AQ): evidence from Asperger syndrome/high-functioning autism, males and females, scientists and mathematicians. J Autism Dev Disord 31(1), 5–17.

Bobo, E., Lin, L., Acquaviva, E., Caci, H., Franc, N., Gamon, L., Picot, M.C., Pupier, F., Speranza, M., Falissard, B., Purper-Ouakil, D., 2020. [How do children and adolescents with Attention Deficit Hyperactivity Disorder (ADHD) experience lockdown during the COVID-19 outbreak?]. Encephale 46(3s), S85–s92.

Boyd, A., Golding, J., Macleod, J., Lawlor, D.A., Fraser, A., Henderson, J., Molloy, L., Ness, A., Ring, S., Davey Smith, G.x, 2013. Cohort Profile: the ‘children of the 90s’--the index offspring of the Avon Longitudinal Study of Parents and Children. Int J Epidemiol 42(1), 111–127.

Bundy, R., Mandy, W., Crane, L., Belcher, H., Bourne, L., Brede, J., Hull, L., Brinkert, J., Cook, J., 2022. The impact of early stages of COVID-19 on the mental health of autistic adults in the United Kingdom: A longitudinal mixed-methods study. Autism, 13623613211065543.

Creswell, C., Shum, A., Pearcey, S., Skripkauskaite, S., Patalay, P., Waite, P., 2021. Young people’s mental health during the COVID-19 pandemic. Lancet Child Adolesc Health 5(8), 535–537.

Davies, S.J.C., Pearson, R.M., Stapinski, L., Bould, H., Christmas, D.M., Button, K.S., Skapinakis, P., Lewis, G., Evans, J., 2016. Symptoms of generalized anxiety disorder but not panic disorder at age 15 years increase the risk of depression at 18 years in the Avon Longitudinal Study of Parents and Children (ALSPAC) cohort study. Psychological medicine 46(1), 73–85.

Douglas, M., Katikireddi, S.V., Taulbut, M., McKee, M., McCartney, G., 2020. Mitigating the wider health effects of covid-19 pandemic response. BMJ 369, m1557.

Eyre, O., Bevan Jones, R., Agha, S.S., Wootton, R.E., Thapar, A.K., Stergiakouli, E., Langley, K., Collishaw, S., Thapar, A., Riglin, L., 2021. Validation of the short Mood and Feelings Questionnaire in young adulthood. J Affect Disord 294, 883–888.

Faraone, S.V., Banaschewski, T., Coghill, D., Zheng, Y., Biederman, J., Bellgrove, M.A., Newcorn, J.H., Gignac, M., Al Saud, N.M., Manor, I., Rohde, L.A., Yang, L., Cortese, S., Almagor, D., Stein, M.A., Albatti, T.H., Aljoudi, H.F., Alqahtani, M.M.J., Asherson, P., Atwoli, L., Bolte, S., Buitelaar, J.K., Crunelle, C.L., Daley, D., Dalsgaard, S., Dopfner, M., Espinet, S., Fitzgerald, M., Franke, B., Gerlach, M., Haavik, J., Hartman, C.A., Hartung, C.M., Hinshaw, S.P., Hoekstra, P.J., Hollis, C., Kollins, S.H., Sandra Kooij, J.J., Kuntsi, J., Larsson, H., Li, T., Liu, J., Merzon, E., Mattingly, G., Mattos, P., McCarthy, S., Mikami, A.Y., Molina, B.S.G., Nigg, J.T., Purper-Ouakil, D., Omigbodun, O.O., Polanczyk, G.V., Pollak, Y., Poulton, A.S., Rajkumar, R.P., Reding, A., Reif, A., Rubia, K., Rucklidge, J., Romanos, M., Ramos-Quiroga, J.A., Schellekens, A., Scheres, A., Schoeman, R., Schweitzer, J.B., Shah, H., Solanto, M.V., Sonuga-Barke, E., Soutullo, C., Steinhausen, H.C., Swanson, J.M., Thapar, A., Tripp, G., van de Glind, G., Brink, W.V.D., Van der Oord, S., Venter, A., Vitiello, B., Walitza, S., Wang, Y., 2021. The World Federation of ADHD International Consensus Statement: 208 Evidence-based conclusions about the disorder. Neurosci Biobehav Rev 128, 789–818.

Farley, M., Cottle, K.J., Bilder, D., Viskochil, J., Coon, H., McMahon, W., 2018. Mid-life social outcomes for a population-based sample of adults with ASD. Autism Res 11(1), 142–152.

Fraser, A., Macdonald-Wallis, C., Tilling, K., Boyd, A., Golding, J., Davey Smith, G., Henderson, J., Macleod, J., Molloy, L., Ness, A., Ring, S., Nelson, S.M., Lawlor, D.A., 2013. Cohort Profile: the Avon Longitudinal Study of Parents and Children: ALSPAC mothers cohort. Int J Epidemiol 42(1), 97–110.

Goodman, R., 1997. The Strengths and Difficulties Questionnaire: A Research Note. Journal of Child Psychology and Psychiatry 38(5), 581–586.

Griffiths, S., Allison, C., Kenny, R., Holt, R., Smith, P., Baron-Cohen, S., 2019. The Vulnerability Experiences Quotient (VEQ): A Study of Vulnerability, Mental Health and Life Satisfaction in Autistic Adults. Autism Res 12(10), 1516–1528.

Gustafsson, M., 2020. Young workers in the coronavirus crisis: Findings from the Resolution Foundation’s coronavirus survey. https://www.resolutionfoundation.org/publications/young-workers-in-the-coronavirus-crisis/. (Accessed 07/04/2022 2022).

Holmes, E.A., O’Connor, R.C., Perry, V.H., Tracey, I., Wessely, S., Arseneault, L., Ballard, C., Christensen, H., Cohen Silver, R., Everall, I., Ford, T., John, A., Kabir, T., King, K., Madan, I., Michie, S., Przybylski, A.K., Shafran, R., Sweeney, A., Worthman, C.M., Yardley, L., Cowan, K., Cope, C., Hotopf, M., Bullmore, E., 2020. Multidisciplinary research priorities for the COVID-19 pandemic: a call for action for mental health science. Lancet Psychiatry 7(6), 547–560.

Houghton, S., Kyron, M., Lawrence, D., Hunter, S.C., Hattie, J., Carroll, A., Zadow, C., Chen, W., 2022. Longitudinal trajectories of mental health and loneliness for Australian adolescents with-or-without neurodevelopmental disorders: the impact of COVID-19 school lockdowns. J Child Psychol Psychiatry.

Hull, L., Lai, M.-C., Baron-Cohen, S., Allison, C., Smith, P., Petrides, K.V., Mandy, W., 2019. Gender differences in self-reported camouflaging in autistic and non-autistic adults. Autism 24(2), 352–363.

Hull, L., Levy, L., Lai, M.-C., Petrides, K.V., Baron-Cohen, S., Allison, C., Smith, P., Mandy, W., 2021. Is social camouflaging associated with anxiety and depression in autistic adults? Molecular Autism 12(1), 13.

Hull, L., Petrides, K.V., Allison, C., Smith, P., Baron-Cohen, S., Lai, M.-C., Mandy, W., 2017. “Putting on My Best Normal”: Social Camouflaging in Adults with Autism Spectrum Conditions. Journal of autism and developmental disorders 47(8), 2519–2534.

Jacobson, N.C., Newman, M.G., 2014. Avoidance mediates the relationship between anxiety and depression over a decade later. J Anxiety Disord 28(5), 437–445.

Johansson, R., Carlbring, P., Heedman, Å., Paxling, B., Andersson, G., 2013. Depression, anxiety and their comorbidity in the Swedish general population: point prevalence and the effect on health-related quality of life. PeerJ 1, e98–e98.

Kaplan, E.L., Meier, P., 1958. Nonparametric Estimation from Incomplete Observations. Journal of the American Statistical Association 53(282), 457–481.

Kwong, A.S.F., Pearson, R.M., Adams, M.J., Northstone, K., Tilling, K., Smith, D., Fawns-Ritchie, C., Bould, H., Warne, N., Zammit, S., Gunnell, D.J., Moran, P.A., Micali, N., Reichenberg, A., Hickman, M., Rai, D., Haworth, S., Campbell, A., Altschul, D., Flaig, R., McIntosh, A.M., Lawlor, D.A., Porteous, D., Timpson, N.J., 2020. Mental health before and during the COVID-19 pandemic in two longitudinal UK population cohorts. Br J Psychiatry, 1–10.

Larsson, H., Anckarsater, H., Råstam, M., Chang, Z., Lichtenstein, P., 2012. Childhood attention-deficit hyperactivity disorder as an extreme of a continuous trait: a quantitative genetic study of 8,500 twin pairs. J Child Psychol Psychiatry 53(1), 73–80.

Lewis, G., Duffy, L., Ades, A., Amos, R., Araya, R., Brabyn, S., Button, K.S., Churchill, R., Derrick, C., Dowrick, C., Gilbody, S., Fawsitt, C., Hollingworth, W., Jones, V., Kendrick, T., Kessler, D., Kounali, D., Khan, N., Lanham, P., Pervin, J., Peters, T.J., Riozzie, D., Salaminios, G., Thomas, L., Welton, N.J., Wiles, N., Woodhouse, R., Lewis, G., 2019. The clinical effectiveness of sertraline in primary care and the role of depression severity and duration (PANDA): a pragmatic, double-blind, placebo-controlled randomised trial. The lancet. Psychiatry 6(11), 903–914.

Lewis, K.J.S., Lewis, C., Roberts, A., Richards, N.A., Evison, C., Pearce, H.A., Lloyd, K., Meudell, A., Edwards, B.M., Robinson, C.A., Poole, R., John, A., Bisson, J.I., Jones, I., 2022. The effect of the COVID-19 pandemic on mental health in individuals with pre-existing mental illness. BJPsych Open 8(2), e59.

Mantel, N., 1966. Evaluation of survival data and two new rank order statistics arising in its consideration. Cancer Chemother Rep 50(3), 163–170.

McGinty, E.E., Presskreischer, R., Anderson, K.E., Han, H., Barry, C.L., 2020a. Psychological Distress and COVID-19–Related Stressors Reported in a Longitudinal Cohort of US Adults in April and July 2020. JAMA 324(24), 2555–2557.

McGinty, E.E., Presskreischer, R., Han, H., Barry, C.L., 2020b. Psychological Distress and Loneliness Reported by US Adults in 2018 and April 2020. JAMA 324(1), 93–94.

Niedzwiedz, C.L., Green, M.J., Benzeval, M., Campbell, D., Craig, P., Demou, E., Leyland, A., Pearce, A., Thomson, R., Whitley, E., Katikireddi, S.V., 2021. Mental health and health behaviours before and during the initial phase of the COVID-19 lockdown: longitudinal analyses of the UK Household Longitudinal Study. Journal of Epidemiology and Community Health 75(3), 224.

Northstone, K., Lewcock, M., Groom, A., Boyd, A., Macleod, J., Timpson, N., Wells, N., 2019. The Avon Longitudinal Study of Parents and Children (ALSPAC): an update on the enrolled sample of index children in 2019. Wellcome Open Res 4, 51.

Oomen, D., Nijhof, A.D., Wiersema, J.R., 2021. The psychological impact of the COVID-19 pandemic on adults with autism: a survey study across three countries. Mol Autism 12(1), 21.

Patel, K., Robertson, E., Kwong, A.S.F., Griffith, G.J., Willan, K., Green, M.J., Gessa, G.D., Huggins, C.F., McElroy, E., Thompson, E.J., Maddock, J., Niedzwiedz, C.L., Henderson, M., Richards, M., Steptoe, A., Ploubidis, G.B., Moltrecht, B., Booth, C., Fitzsimons, E., Silverwood, R., Patalay, P., Porteous, D., Katikireddi, S.V., 2021. Psychological Distress Before and During the COVID-19 Pandemic: Sociodemographic Inequalities in 11 UK Longitudinal Studies. medRxiv, 2021.2010.2022.21265368.

Pierce, M., McManus, S., Hope, H., Hotopf, M., Ford, T., Hatch, S.L., John, A., Kontopantelis, E., Webb, R.T., Wessely, S., Abel, K.M., 2021. Mental health responses to the COVID-19 pandemic: a latent class trajectory analysis using longitudinal UK data. The Lancet Psychiatry 8(7), 610–619.

Pierre, M., Keller, M., Altschul, D., Fawns-Ritchie, C., Hartley, L., Nangle, C., Edwards, R., Dawson, R., Campbell, A., Flaig, R., Porteous, D., 2021. Socioeconomic position and mental health during the COVID-19 pandemic: a cross-sectional analysis of the CovidLife study [version 1; peer review: awaiting peer review]. Wellcome Open Research 6(139).

Pijls, B.G., Jolani, S., Atherley, A., Derckx, R.T., Dijkstra, J.I.R., Franssen, G.H.L., Hendriks, S., Richters, A., Venemans-Jellema, A., Zalpuri, S., Zeegers, M.P., 2021. Demographic risk factors for COVID-19 infection, severity, ICU admission and death: a meta-analysis of 59 studies. BMJ Open 11(1), e044640.

R Core Team, 2021. R: A Language and Environment for Statistical Computing. R Foundation for Statistical Computing, Vienna, Austria.

Riglin, L., Agha, S.S., Eyre, O., Bevan Jones, R., Wootton, R.E., Thapar, A.K., Collishaw, S., Stergiakouli, E., Langley, K., Thapar, A., 2021a. Investigating the validity of the Strengths and Difficulties Questionnaire to assess ADHD in young adulthood. Psychiatry Res 301, 113984.

Riglin, L., Leppert, B., Dardani, C., Thapar, A.K., Rice, F., O’Donovan, M.C., Davey Smith, G., Stergiakouli, E., Tilling, K., Thapar, A., 2021b. ADHD and depression: investigating a causal explanation. Psychol Med 51(11), 1890–1897.

Riglin, L., Todd, A., Blakey, R., Shakeshaft, A., Stergiakouli, E., Davey Smith, G., Tilling, K., Thapar, A., in press. Investigating young-adult social outcomes of attention deficit hyperactivity disorder. The Journal of Clinical Psychiatry.

Roberts, D.L., Rossman, J.S., Jaric, I., 2021. Dating first cases of COVID-19. PLoS Pathog 17(6), e1009620.

Robinson, E., Sutin, A.R., Daly, M., Jones, A., 2022. A systematic review and meta-analysis of longitudinal cohort studies comparing mental health before versus during the COVID-19 pandemic in 2020. Journal of affective disorders 296, 567–576.

Rutter, M., Pine, D.S., 2015. Diagnosis, diagnostic formulations, and classification, Rutter’s Child and Adolescent Psychiatry. pp. 17–30.

Santomauro, D.F., Mantilla Herrera, A.M., Shadid, J., Zheng, P., Ashbaugh, C., Pigott, D.M., Abbafati, C., Adolph, C., Amlag, J.O., Aravkin, A.Y., Bang-Jensen, B.L., Bertolacci, G.J., Bloom, S.S., Castellano, R., Castro, E., Chakrabarti, S., Chattopadhyay, J., Cogen, R.M., Collins, J.K., Dai, X., Dangel, W.J., Dapper, C., Deen, A., Erickson, M., Ewald, S.B., Flaxman, A.D., Frostad, J.J., Fullman, N., Giles, J.R., Giref, A.Z., Guo, G., He, J., Helak, M., Hulland, E.N., Idrisov, B., Lindstrom, A., Linebarger, E., Lotufo, P.A., Lozano, R., Magistro, B., Malta, D.C., Månsson, J.C., Marinho, F., Mokdad, A.H., Monasta, L., Naik, P., Nomura, S., O’Halloran, J.K., Ostroff, S.M., Pasovic, M., Penberthy, L., Reiner Jr, R.C., Reinke, G., Ribeiro, A.L.P., Sholokhov, A., Sorensen, R.J.D., Varavikova, E., Vo, A.T., Walcott, R., Watson, S., Wiysonge, C.S., Zigler, B., Hay, S.I., Vos, T., Murray, C.J.L., Whiteford, H.A., Ferrari, A.J., 2021. Global prevalence and burden of depressive and anxiety disorders in 204 countries and territories in 2020 due to the COVID-19 pandemic. The Lancet 398(10312), 1700–1712.

Spitzer, R.L., Kroenke, K., Williams, J.B., Lowe, B., 2006. A brief measure for assessing generalized anxiety disorder: the GAD-7. Arch Intern Med 166(10), 1092–1097.

Taylor, A.E., Jones, H.J., Sallis, H., Euesden, J., Stergiakouli, E., Davies, N.M., Zammit, S., Lawlor, D.A., Munafo, M.R., Davey Smith, G., Tilling, K., 2018. Exploring the association of genetic factors with participation in the Avon Longitudinal Study of Parents and Children. Int J Epidemiol 47(4), 1207–1216.

Tennant, R., Hiller, L., Fishwick, R., Platt, S., Joseph, S., Weich, S., Parkinson, J., Secker, J., Stewart-Brown, S., 2007. The Warwick-Edinburgh Mental Well-being Scale (WEMWBS): development and UK validation. Health Qual Life Outcomes 5, 63.

Thapar, A., Cooper, M., 2016. Attention deficit hyperactivity disorder. The Lancet 387(10024), 1240–1250.

Vadivel, R., Shoib, S., El Halabi, S., El Hayek, S., Essam, L., Gashi Bytyci, D., Karaliuniene, R., Schuh Teixeira, A.L., Nagendrappa, S., Ramalho, R., Ransing, R., Pereira-Sanchez, V., Jatchavala, C., Adiukwu, F.N., Kudva Kundadak, G., 2021. Mental health in the post-COVID-19 era: challenges and the way forward. Gen Psychiatr 34(1), e100424.

Vasa, R.A., Singh, V., Holingue, C., Kalb, L.G., Jang, Y., Keefer, A., 2021. Psychiatric problems during the COVID-19 pandemic in children with autism spectrum disorder. Autism Res, 10.1002/aur.2574.

Warne, N., Heron, J., Mars, B., Kwong, A.S.F., Solmi, F., Pearson, R., Moran, P., Bould, H., 2021. Disordered eating and self-harm as risk factors for poorer mental health during the COVID-19 pandemic: a UK-based birth cohort study. Journal of Eating Disorders 9(1), 155.

Warwick Medical School., 2021. Collect, score, analyse and interpret WEMWBS. https://warwick.ac.uk/fac/sci/med/research/platform/wemwbs/using/howto. (Accessed 1/11/21.

Zhang, Z., Parker, R.M.A., Charlton, C.M.J., Leckie, G., Browne, W.J., 2016. R2MLwiN: A Package to Run MLwiN from within R. Journal of Statistical Software 72(10), 1–43.

