## Supplementary for "Mental-health before and during the COVID-19 pandemic in adults with neurodevelopmental disorders"

### **Supplementary Material**

#### **ALSPAC study information**

Pregnant women resident in Avon, UK with expected dates of delivery 1st April 1991 to 31st December 1992 were invited to take part in the study. The initial number of pregnancies enrolled is 14,541 (for these at least one questionnaire has been returned or a “Children in Focus” clinic had been attended by 19/07/99). Of these initial pregnancies, there was a total of 14,676 foetuses, resulting in 14,062 live births and 13,988 children who were alive at 1 year of age.

When the oldest children were approximately 7 years of age, an attempt was made to bolster the initial sample with eligible cases who had failed to join the study originally. As a result, when considering variables collected from the age of seven onwards (and potentially abstracted from obstetric notes) there are data available for more than the 14,541 pregnancies mentioned above. The number of **new pregnancies** not in the initial sample (known as Phase I enrolment) that are currently represented on the built files and reflecting enrolment status at the age of 24 is 913 (456, 262 and 195 recruited during Phases II, III and IV respectively), resulting in an additional 913 children being enrolled. The phases of enrolment are described in more detail in the cohort profile paper and its update (see footnote 4 below). The total sample size for analyses using any data collected after the age of seven is therefore 15,454 pregnancies, resulting in 15,589 foetuses. Of these 14,901 were **alive at 1 year of age**.

A 10% sample of the ALSPAC cohort, known as the **Children in Focus (CiF) group**, attended clinics at the University of Bristol at various time intervals between 4 to 61 months of age. The CiF group were chosen at random from the last 6 months of ALSPAC births (1432 families attended at least one clinic). Excluded were those mothers who had moved out of the area or were lost to follow-up, and those partaking in another study of infant development in Avon.

Study data were collected and managed using REDCap electronic data capture tools hosted at the University of Bristol (Harris et al., 2009). REDCap (Research Electronic Data Capture) is a secure, web-based software platform designed to support data capture for research studies. Ethical approval for the study was obtained from the ALSPAC Ethics and Law Committee and the Local Research Ethics Committees. Informed consent for the use of data collected via questionnaires and clinics was obtained from participants following the recommendations of the ALSPAC Ethics and Law Committee at the time.

The study website contains details of all the data that is available through a fully searchable data dictionary and variable search tool (<http://www.bristol.ac.uk/alspac/researchers/our-data>/).

#### **Attrition and missing data**

The primary sample for analysis of mental health consisted of those with complete neurodevelopmental (AQ & SDQ), sex and COVID-19 event data (see main methods) and mental health outcome data from at least one timepoint (pre-pandemic, COVID1, COVID2 or COVID3) (n=3058). For anxiety, 1997 (65%) had GAD7 data at age 21, 2048 (67%) at COVID1, 1897 (62%) at COVID2 and 3028 (99%) at COVID3. For depression, 2973 (97%) had SMFQ data at age 25, 2024 (66%) at COVID1, 1892 (62%) at COVID2 and 2997 (98%) at COVID3. For mental wellbeing, 2192 (72%) had WEMWBS data at age 23, 2035 (67%) at COVID1, 1902 (62%) at COVID2 and 3014 (99%) at COVID3.

#### **Imputation of missing questionnaire items**

SDQ hyperactivity/inattentive questionnaires were discarded where >2 items were missing. In line with recommendations ([www.sdqinfo.org](http://www.sdqinfo.org)) total subscale scores were derived using mean imputation for those with (≤2) of items missing.

AQ-10 and AQ-28 scores were only calculated when all items were completed. For the 50-item AQ, questionnaires with >10% of items missing were not used. Where between 1-5 items were missing, items were corrected as follows: corrected score=raw score x (50 / (50 – missing items).

#### **Correlation between mental health scores**

Correlations between continuous GAD7, SMFQ and WEMWBS scores at each timepoint are presented in Supplementary Table 8.

#### Using broader definitions of ASD and ADHD sensitivity analysis

We repeated the main analysis of mental health using broader definitons of ADHD (SDQ >= 5) and ASD (AQ >27). Using these cut-offs, 24% of the cohort was categorised as having broad ADHD and 11% categorised as broad ASD. The cohort information remained similar in Supplementary Table 7 to the original cut-offs in Table 1, with the only differences being a higher proportion of individuals with broad ADHD reporting losing their jobs during the pandemic than those without, and a higher proportion of individuals with broad ASD reporting financial difficulties during the pandemic, differences we did not see using the original cut-points.

Multilevel analysis using the broader cut-points for ADHD showed very similar β coefficients for fixed effects as using the original cut-point (estimates presented for ADHD in Supplementary Figure 5 and for ASD in Supplementary Figure 6).

#### **COVID-19 infection**

Data on COVID-19 infection up to the third COVID data sweep (Jan-March 2021) in ALSPAC was available in 3,124 people with complete ADHD and ASD symptom scores. 15.8% (493/3124) of the total cohort, encompassing 14% (55/394) of those with ADHD and 10% (8/79) of those with ASD reported a COVID-19 infection between the start of the pandemic and the time of COVID3 survey completion (Jan-March 2021). There was not strong evidence of association between reporting a COVID-19 infection and being classified as ADHD (OR = 0.85 [0.62-1.14], p=0.29) or ASD (OR = 0.61 [0.27-1.19], p=0.16).

A log rank test was run to determine if there were differences in the distribution of infections in those with ADHD and ASD compared to those without. There was not strong evidence of differences in infection incidence distribution for either subgroup (ADHD (*χ*^2^(1)=1.1, p=0.29; ASD (*χ*^2^(1)=1.7, p=0.19). Cumulative incidence curves are presented in Supplementary Figure 7.

#### **References**

Harris, P.A., Taylor, R., Thielke, R., Payne, J., Gonzalez, N., Conde, J.G., 2009. Research electronic data capture (REDCap)--a metadata-driven methodology and workflow process for providing translational research informatics support. J Biomed Inform 42(2), 377-381.

#### **Supplementary figures**

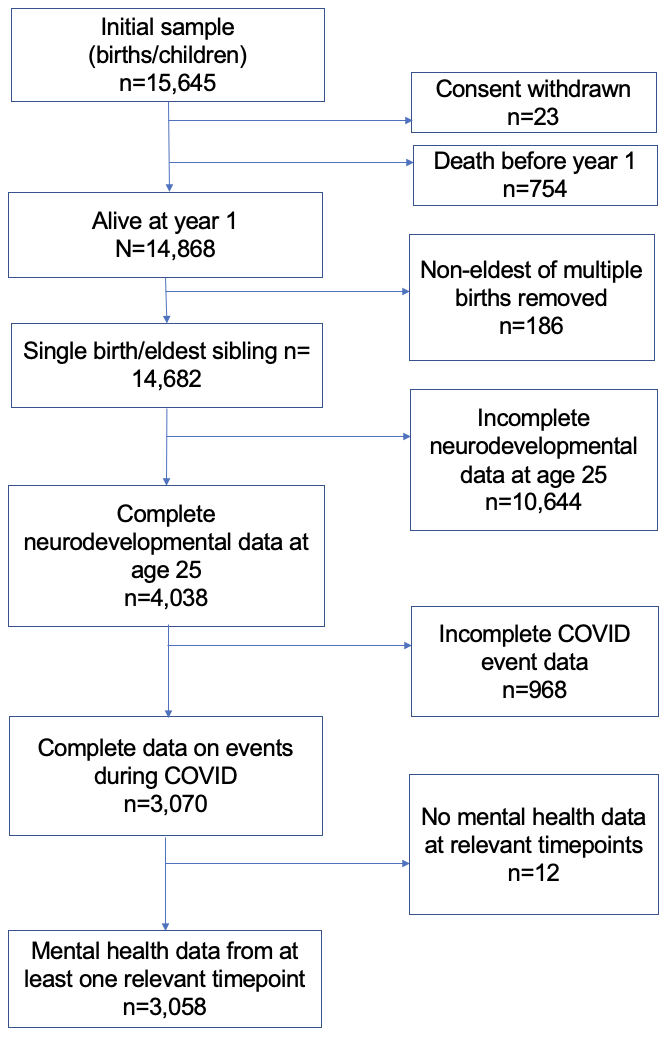

**Supplementary Figure 1** - *Details of sample used for analysis of mental health during COVID-19 pandemic.*

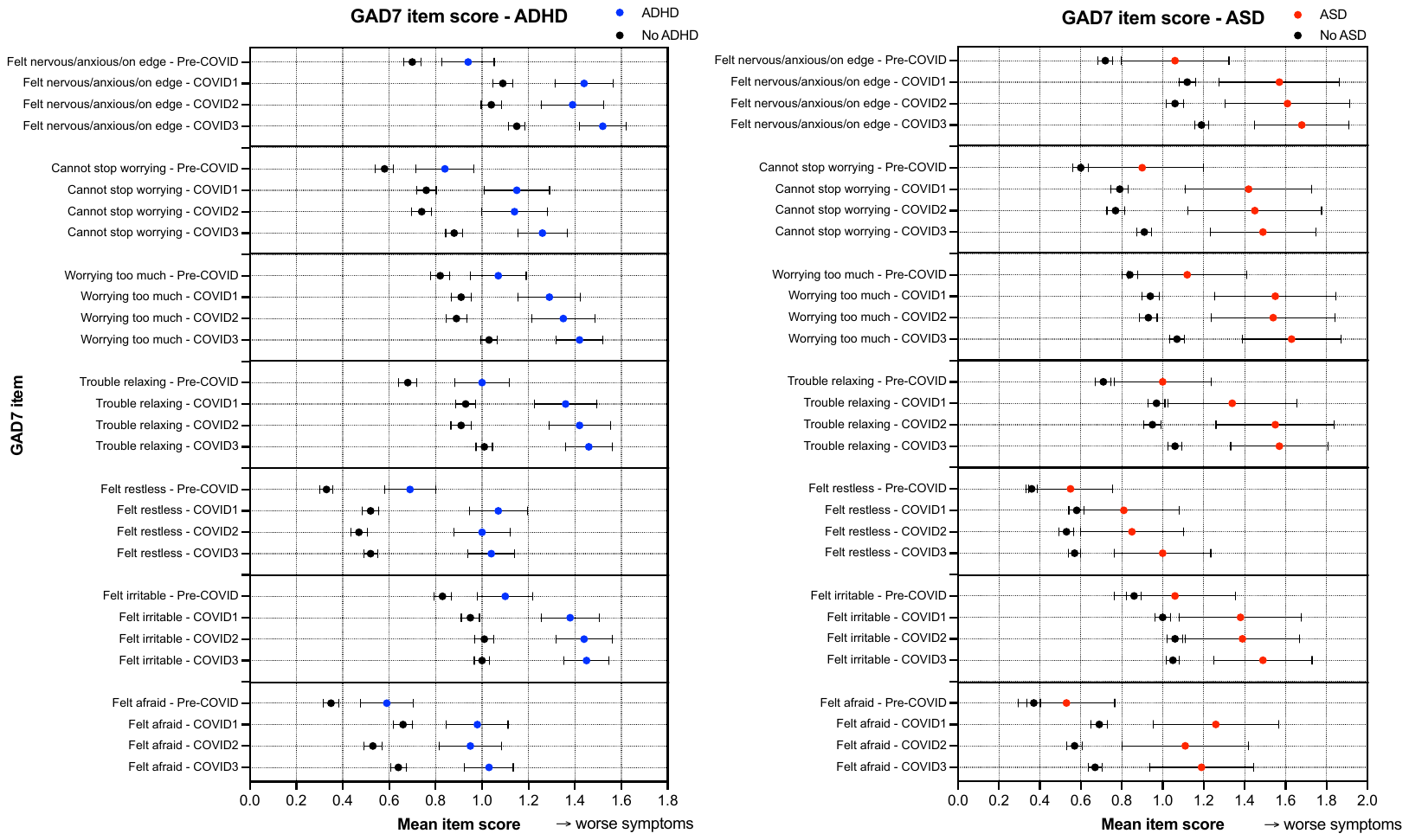

**Supplementary Figure 2** - *Item by item comparison of GAD7 questionnaires in those with and without ADHD/ASD at each questionnaire timepoint. Mean scores ± 95% confidence intervals are plotted. All items indicated worse symptoms during COVID timepoints, for all subgroups.*

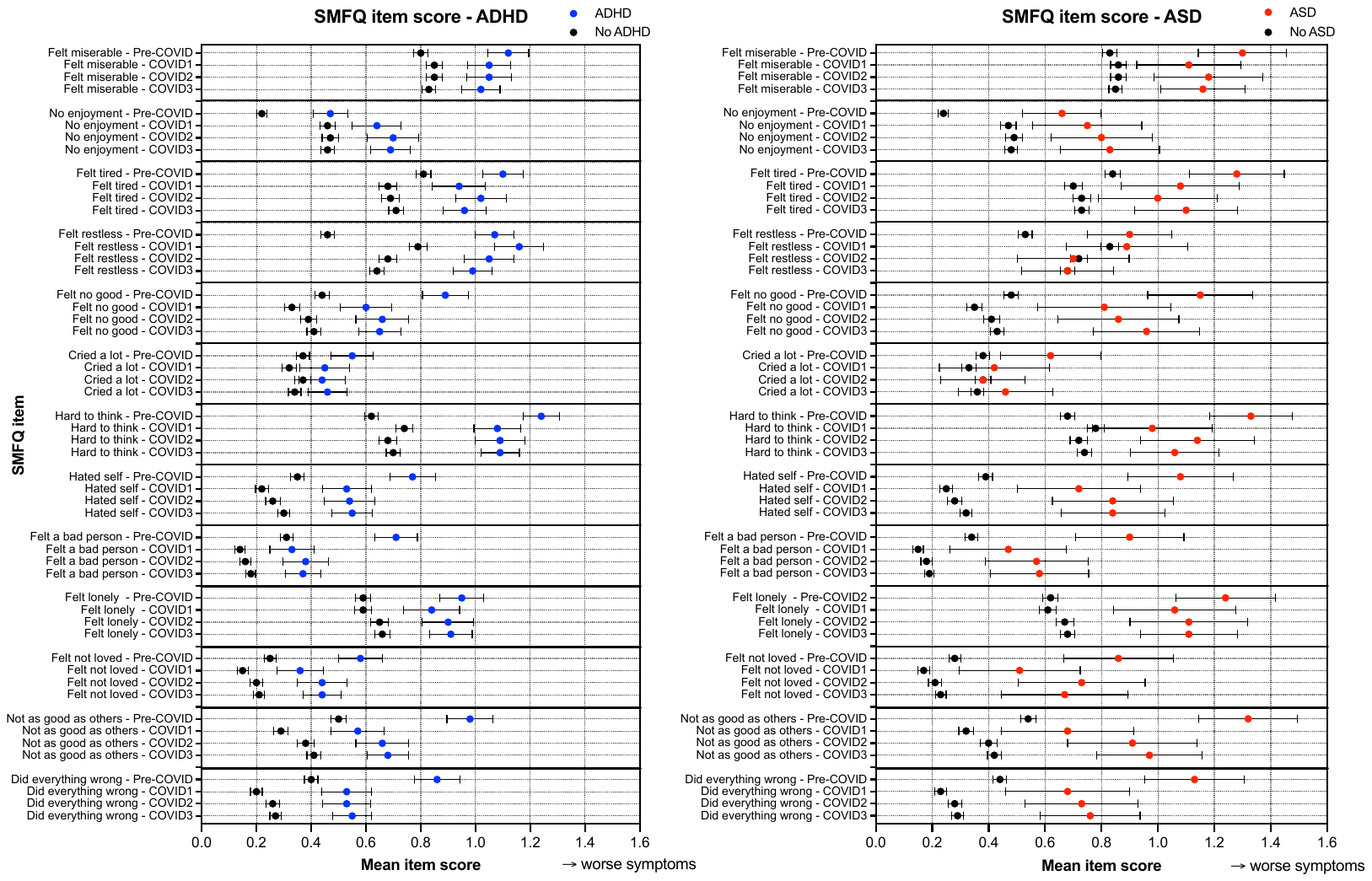

**Supplementary Figure 3** - *Item by item comparison of SMFQ questionnaires in those with and without ADHD/ASD at each questionnaire timepoint. Mean scores ± 95% confidence intervals are plotted. Almost all items (except ‘no enjoyment’) indicated better symptoms during COVID timepoints compared to pre-pandemic in all subgroups. Feeling lonely was marginally elevated at COVID2 and 3 timepoints in those with ASD and ADHD at a level higher than pre-pandemic, whereas remained lower than pre-pandemic levels in those with ADHD and ASD.*

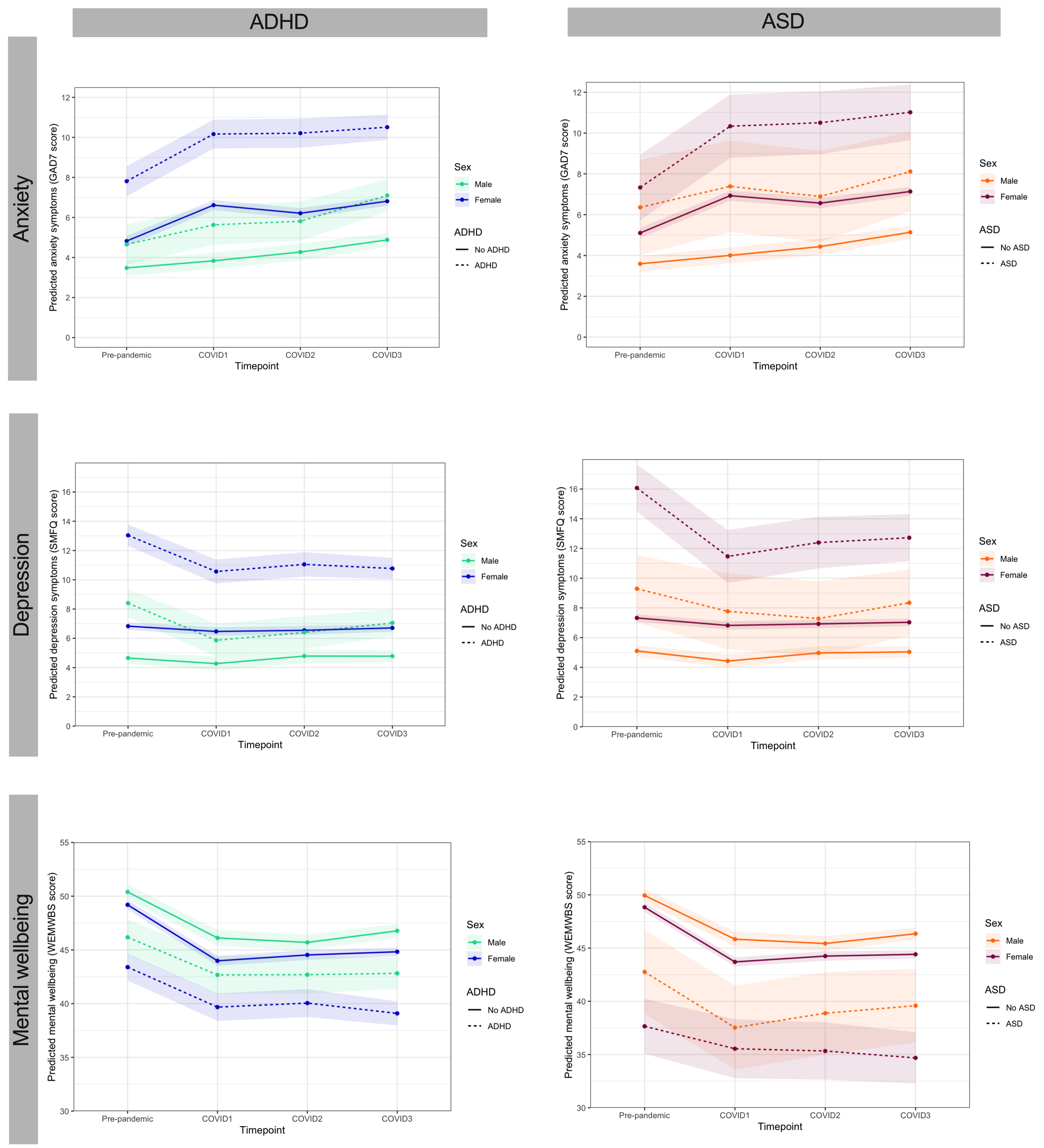

**Supplementary Figure 4** - *Trajectories of mental health before and during the COVID-19 pandemic, stratified by sex and ADHD/ASD status.*

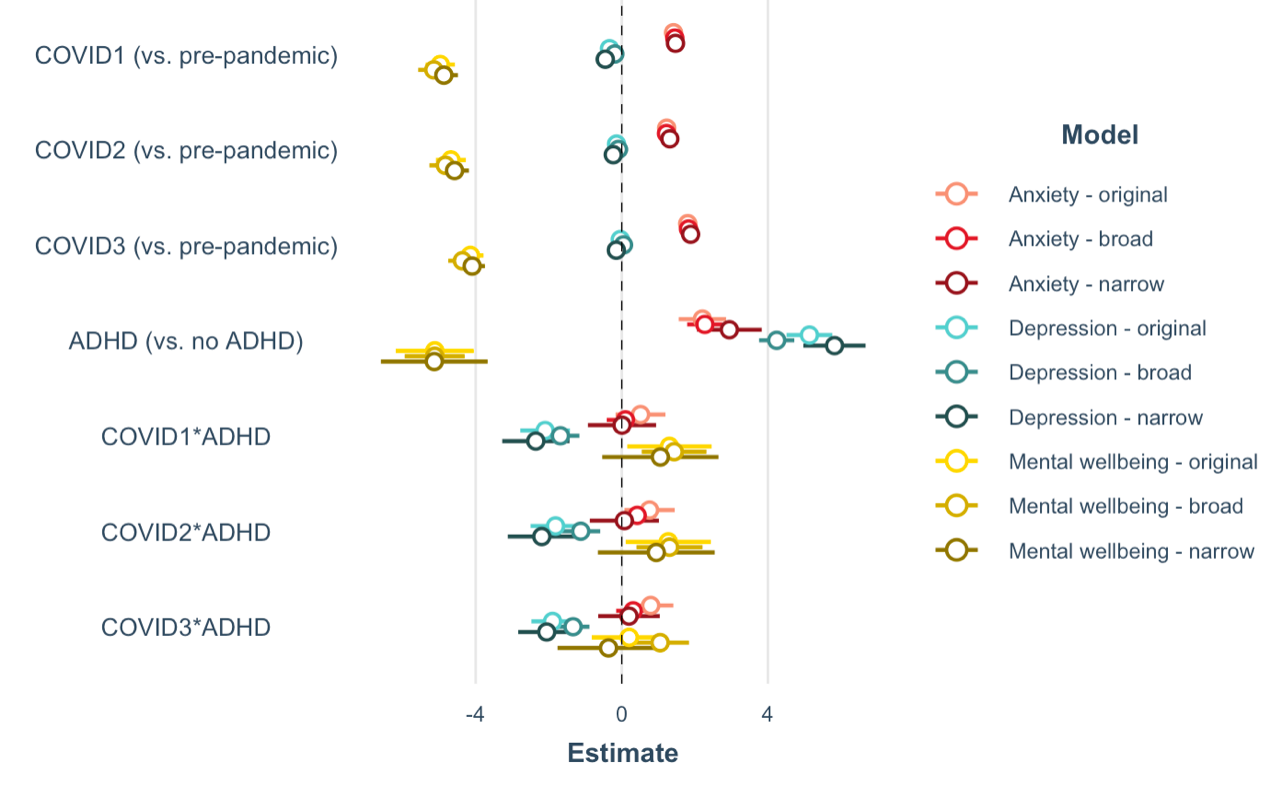

**Supplementary Figure 5** - *Beta coefficients from multilevel models of mental health outcomes in ADHD, using the original SDQ cut-point of 6 and the broader cut-point of 5. Estimates are very similar to in using both definitions, just with a weaker effect for broader definitions.*

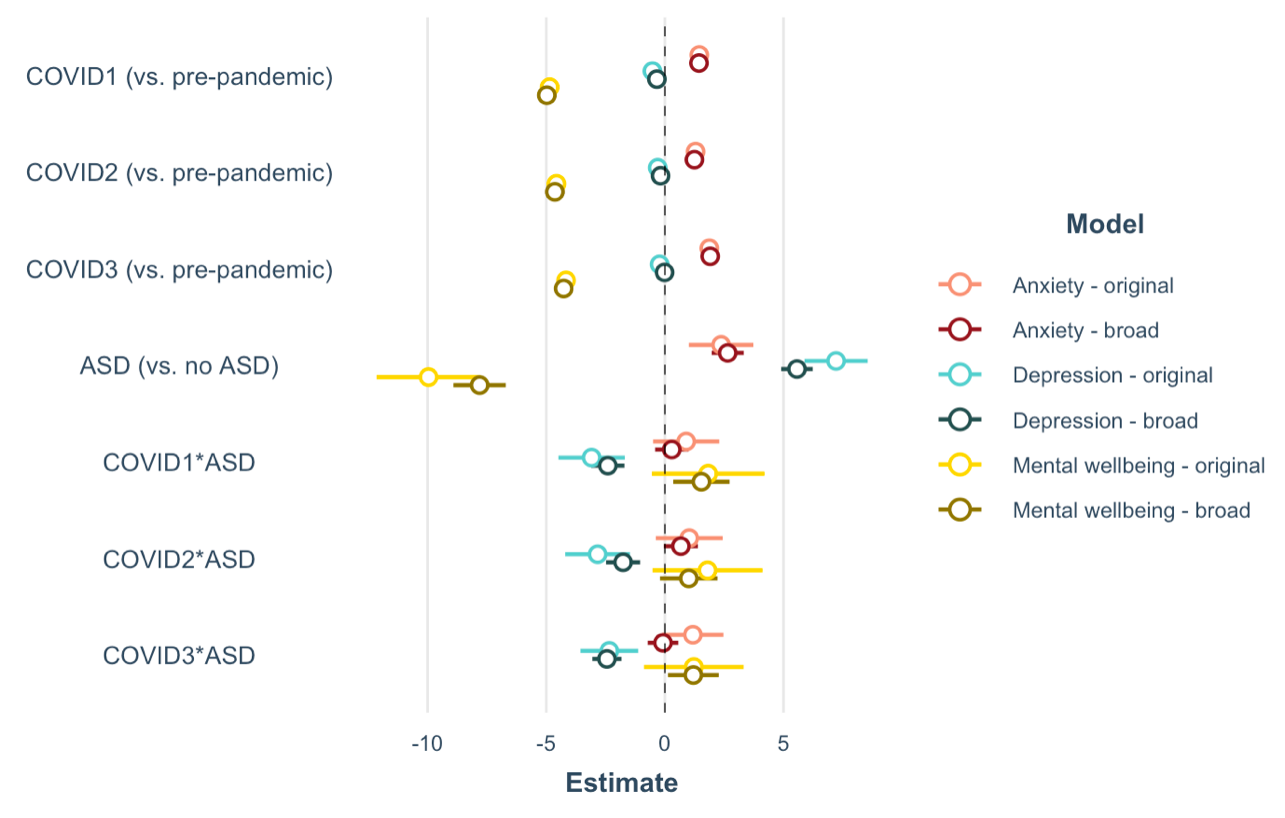

**Supplementary Figure 6** - *Beta coefficients from multilevel models of mental health outcomes in ASD, using the original SDQ cut-point of 6 and the broader cut-point of 5. Estimates are very similar to in using both definitions, just with a weaker effect for broader definitions.*

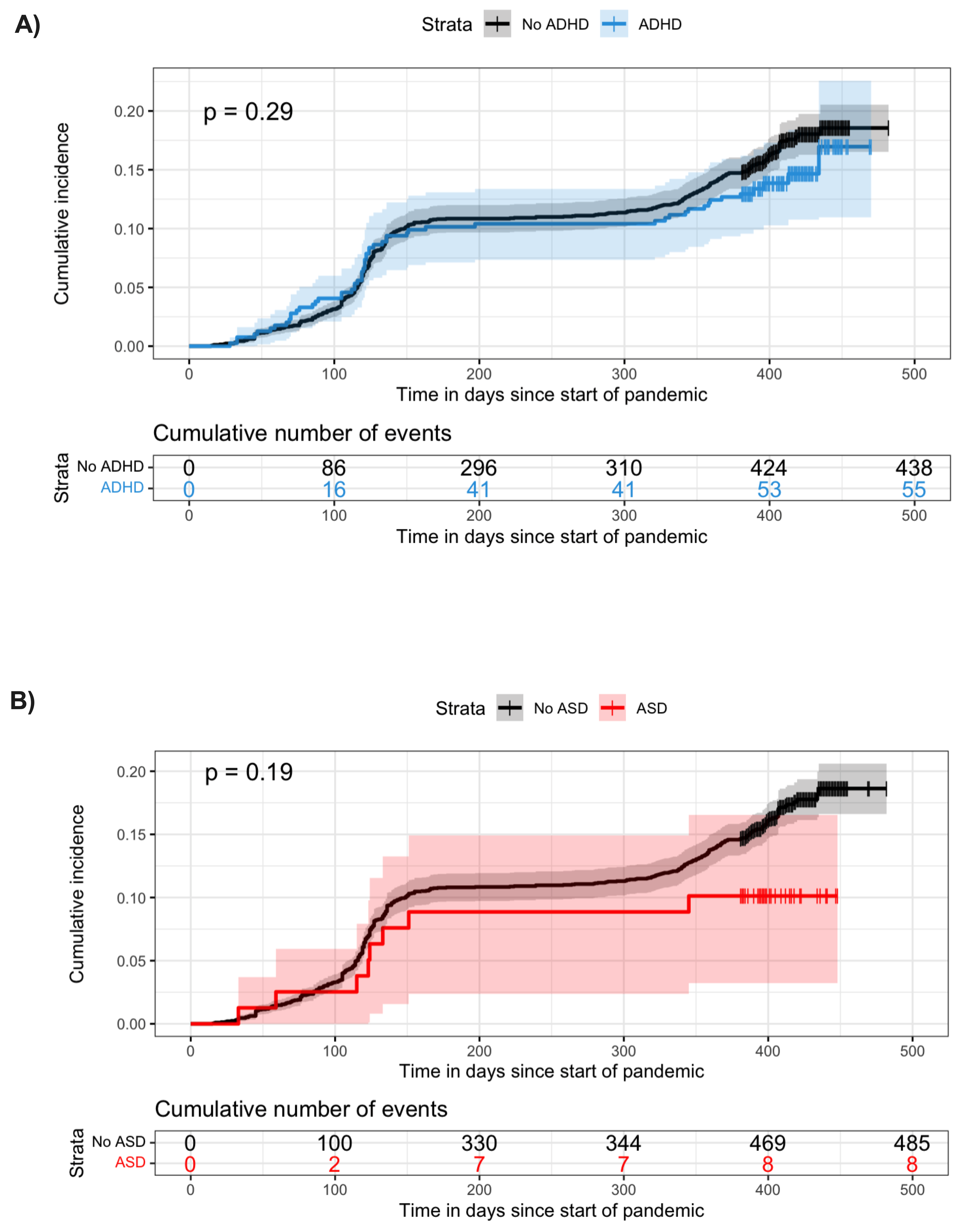

**Supplementary Figure 7** - *Survival plots showing cumulative incidence of COVID-19 infection in those with and without ADHD (A) and ASD (B).*

#### **Supplementary tables**

**Supplementary Table 1** - Multilevel model of anxiety and ADHD. Beta coefficients [95% CIs] are presented for each variable in the four models. Positive beta coefficients indicate worse anxiety symptoms.

| **Anxiety** | **Model 1**  Total | | **Model 2**  Male strata | | | **Model 3**  Female strata | | | **Model 4**  Total, sex-interaction | |
| --- | --- | --- | --- | --- | --- | --- | --- | --- | --- | --- |
|  | Beta coefficient  [95% CIs] | P-value | Beta coefficient  [95% CIs] | P-value | Beta coefficient  [95% CIs] | | P-value | Beta coefficient  [95% CIs] | | P-value |
| **Fixed effects** | | | | | | | | | | |
| *Intercept* | *4.42* | *<2.2x10^-300^* | *3.48* | *4.9x10^-86^* | *4.82* | | *5.1x10^-265^* | *3.48* | | *1.8x10^-66^* |
|  | *[4.20, 4.64]* |  | *[3.13, 3.83]* |  | *[4.55, 5.09]* | |  | *[3.08, 3.88]* | |  |
| COVID1 | 1.41 | **2.8x10^-33^** | 0.36 | **0.07** | 1.79 | | **1.6x10^-35^** | 0.35 | | 0.11 |
|  | [1.18, 1.65] |  | [-0.03, 0.74] |  | [1.51, 2.08] | |  | [-0.08, 0.79] | |  |
| COVID2 | 1.23 | **2.1x10^-24^** | 0.79 | **6.9x10^-5^** | 1.39 | | **6.5x10^-21^** | 0.79 | | **0.0005** |
|  | [0.99, 1.46] |  | [0.40, 1.18] |  | [1.10, 1.68] | |  | [0.35, 1.23] | |  |
| COVID3 | 1.80 | **1.6x10^-63^** | 1.40 | **3.8x10^-16^** | 1.99 | | **2.4x10^-50^** | 1.40 | | **7.4x10^-13^** |
|  | [1.59, 2.01] |  | [1.06, 1.74] |  | [1.73, 2.25] | |  | [1.02, 1.78] | |  |
| ADHD | 2.21 | **2.6x10^-11^** | 1.17 | **0.01** | 2.99 | | **2.0x10^-12^** | 1.17 | | **0.03** |
|  | [1.56, 2.85] |  | [0.24, 2.11] |  | [2.16, 3.82] | |  | [0.11, 2.24] | |  |
| **COVID1*ADHD** | 0.51 | 0.14 | 0.62 | 0.25 | 0.56 | | 0.21 | 0.62 | | 0.31 |
|  | [-0.17, 1.20] |  | [-0.43, 1.67] |  | [-0.31, 1.43] | |  | [-0.57, 1.81] | |  |
| **COVID2*ADHD** | 0.76 | **0.03** | 0.37 | 0.48 | 1.01 | | **0.03** | 0.37 | | 0.53 |
|  | [0.07, 1.45] |  | [-0.67, 1.41] |  | [0.13, 1.89] | |  | [-0.81, 1.55] | |  |
| **COVID3*ADHD** | 0.79 | **0.01** | 1.05 | **0.02** | 0.71 | | **0.08** | 1.05 | | **0.05** |
|  | [0.17, 1.41] |  | [0.13, 1.96] |  | [-0.10, 1.51] | |  | [0.01, 2.09] | |  |
| Sex |  |  |  |  |  | |  | 1.34 | | **2.7x10^-8^** |
|  |  |  |  |  |  | |  | [0.87, 1.82] | |  |
| COVID1*Sex |  |  |  |  |  | |  | 1.44 | | **3.8x10^-8^** |
|  |  |  |  |  |  | |  | [0.93, 1.95] | |  |
| COVID2*Sex |  |  |  |  |  | |  | 0.60 | | **0.03** |
|  |  |  |  |  |  | |  | [0.07, 1.12] | |  |
| COVID3*Sex |  |  |  |  |  | |  | 0.59 | | **0.01** |
|  |  |  |  |  |  | |  | [0.13, 1.05] | |  |
| ADHD*Sex |  |  |  |  |  | |  | 1.82 | | **0.008** |
|  |  |  |  |  |  | |  | [0.48, 3.15] | |  |
| COVID1*ADHD*Sex |  |  |  |  |  | |  | -0.06 | | 0.93 |
|  |  |  |  |  |  | |  | [-1.51, 1.39] | |  |
| COVID2*ADHD*Sex |  |  |  |  |  | |  | 0.64 | | 0.39 |
|  |  |  |  |  |  | |  | [-0.81, 2.09] | |  |
| COVID3*ADHD*Sex |  |  |  |  |  | |  | -0.34 | | 0.61 |
|  |  |  |  |  |  | |  | [-1.63, 0.95] | |  |
| **Random effects** | | | | | | | | | | |
| *Individual - Intercept* | 14.73 | **1.0x10^-196^** | 10.28 | **9.6x10^-58^** | 15.18 | | **6.1x10^-135^** | 13.66 | | **2.4x10^-190^** |
|  | [13.77, 15.70] |  | [9.02, 11.53] |  | [13.97, 16.38] | |  | [12.75, 14.57] | |  |
| *Timepoint - Intercept* | 11.33 | **<2.2x10^-300^** | 8.75 | **6.3x10^-188^** | 12.27 | | **<2.2x10^-300^** | 11.25 | | **<2.2x10^-300^** |
|  | [10.92, 11.73] |  | [8.17, 9.34] |  | [11.75, 12.79] | |  | [10.85, 11.65] | |  |
| **Model information** | | | | | | | | | | |
| N observations | 8971 | | 2653 | | | 6318 | | | 8971 | |
| logLik | -25944.58 | |  | | |  | | | -25833.79 | |
| AIC | 51909.15 | |  | | |  | | | 51703.58 | |
| BIC | 51980.17 | |  | | |  | | | 51831.41 | |

**Supplementary Table 2** - *Multilevel model of anxiety and ASD. Beta coefficients [95% CIs] are presented for each variable in the four models. Positive beta coefficients indicate worse anxiety symptoms.*

| **Anxiety** | **Model 1**  Total | | **Model 2**  Male strata | | | **Model 3**  Female strata | | | **Model 4**  Total, sex-interaction | |
| --- | --- | --- | --- | --- | --- | --- | --- | --- | --- | --- |
|  | Beta coefficient  [95% CIs] | P-value | Beta coefficient  [95% CIs] | P-value | Beta coefficient  [95% CIs] | | P-value | Beta coefficient  [95% CIs] | | P-value |
| **Fixed effects** | | | | | | | | | | |
| *Intercept* | *4.64* | *<2.2x10^-300^* | *3.59* | *2.6x10^-102^* | *5.11* | | *<2.2x10^-300^* | *3.59* | | *5.8x10^-78^* |
|  | *[4.43, 4.85]* |  | *[3.26, 3.92]* |  | *[4.84, 5.37]* | |  | *[3.22, 3.97]* | |  |
| COVID1 | 1.45 | **3.9x10^-38^** | 0.42 | **0.02** | 1.83 | | **6.6x10^-40^** | 0.41 | | **0.05** |
|  | [1.23, 1.67] |  | [0.05, 0.78] |  | [1.56, 2.10] | |  | [0.00, 0.82] | |  |
| COVID2 | 1.29 | **1.5x10^-29^** | 0.85 | **6.3x10^-6^** | 1.46 | | **5.5x10^-25^** | 0.84 | | **7.1x10^-5^** |
|  | [1.07, 1.52] |  | [0.48, 1.21] |  | [1.18, 1.74] | |  | [0.43, 1.26] | |  |
| COVID3 | 1.87 | **5.0x10^-75^** | 1.55 | **1.5x10^-21^** | 2.03 | | **6.7x10^-57^** | 1.54 | | **5.0x10^-17^** |
|  | [1.67, 2.07] |  | [1.23, 1.86] |  | [1.78, 2.28] | |  | [1.18, 1.90] | |  |
| ASD | 2.37 | **0.0006** | 2.75 | **0.009** | 2.23 | | **0.01** | 2.77 | | **0.02** |
|  | [1.01, 3.73] |  | [0.69, 4.81] |  | [0.52, 3.94] | |  | [0.40, 5.13] | |  |
| **COVID1*ASD** | 0.90 | 0.21 | 0.62 | 0.58 | 1.17 | | 0.19 | 0.62 | | 0.62 |
|  | [-0.50, 2.29] |  | [-1.55, 2.79] |  | [-0.59, 2.93] | |  | [-1.84, 3.07] | |  |
| **COVID2*ASD** | 1.03 | 0.15 | -0.28 | 0.80 | 1.71 | | 0.06 | -0.31 | | 0.81 |
|  | [-0.39, 2.44] |  | [-2.50, 1.93] |  | [-0.07, 3.48] | |  | [-2.82, 2.20] | |  |
| **COVID3*ASD** | 1.18 | 0.07 | 0.23 | 0.82 | 1.65 | | **0.05** | 0.21 | | 0.85 |
|  | [-0.11, 2.47] |  | [-1.77, 2.22] |  | [0.03, 3.27] | |  | [-2.05, 2.48] | |  |
| Sex |  |  |  |  |  | |  | 1.51 | | **6.2x10^-11^** |
|  |  |  |  |  |  | |  | [1.06, 1.96] | |  |
| COVID1*Sex |  |  |  |  |  | |  | 1.42 | | **1.2x10^-8^** |
|  |  |  |  |  |  | |  | [0.93, 1.90] | |  |
| COVID2*Sex |  |  |  |  |  | |  | 0.62 | | **0.01** |
|  |  |  |  |  |  | |  | [0.13, 1.11] | |  |
| COVID3*Sex |  |  |  |  |  | |  | 0.49 | | **0.03** |
|  |  |  |  |  |  | |  | [0.06, 0.92] | |  |
| ASD*Sex |  |  |  |  |  | |  | -0.54 | | 0.71 |
|  |  |  |  |  |  | |  | [-3.41, 2.33] | |  |
| COVID1*ASD*Sex |  |  |  |  |  | |  | 0.56 | | 0.71 |
|  |  |  |  |  |  | |  | [-2.42, 3.54] | |  |
| COVID2*ASD*Sex |  |  |  |  |  | |  | 2.02 | | 0.19 |
|  |  |  |  |  |  | |  | [-1.01, 5.05] | |  |
| COVID3*ASD*Sex |  |  |  |  |  | |  | 1.44 | | 0.30 |
|  |  |  |  |  |  | |  | [-1.30, 4.19] | |  |
| **Random effects** | | | | | | | | | | |
| *Individual - Intercept* | 15.32 | **4.1x10^-200^** | 10.48 | **3.3x10^-58^** | 16.15 | | **7.9x10^-139^** | 14.40 | | **4.1x10^-195^** |
|  | [14.32, 16.31] |  | [9.20, 11.76] |  | [14.89, 17.42] | |  | [13.45, 15.35] | |  |
| *Timepoint - Intercept* | 11.33 | **<2.2x10^-300^** | 8.77 | **5.5x10^-188^** | 12.28 | | **<2.2x10^-300^** | 11.26 | | **<2.2x10^-300^** |
|  | [10.92, 11.74] |  | [8.18, 9.36] |  | [11.76, 12.80] | |  | [10.85, 11.66] | |  |
| **Model information** | | | | | | | | | | |
| N | 8971 | | 2653 | | | 6318 | | | 8971 | |
| logLik | -25991.92 | |  | | |  | | | -25898.07 | |
| AIC | 52003.84 | |  | | |  | | | 51832.14 | |
| BIC | 52074.86 | |  | | |  | | | 51959.97 | |

**Supplementary Table 3** - *Multilevel model of depression and ADHD. Beta coefficients [95% CIs] are presented for each variable in the four models. Positive beta coefficients indicate worse depressive symptoms.*

| **Depression** | **Model 1**  Total | | **Model 2**  Male strata | | **Model 3**  Female strata | | **Model 4**  Total, sex-interaction | |
| --- | --- | --- | --- | --- | --- | --- | --- | --- |
|  | Beta coefficient  [95% CIs] | P-value | Beta coefficient  [95% CIs] | P-value | Beta coefficient  [95% CIs] | P-value | Beta coefficient  [95% CIs] | P-value |
| **Fixed effects** | | | | | | | | |
| *Intercept* | *6.17* | *<2.2x10^-300^* | *4.65* | *6.4x10^-157^* | *6.83* | *<2.2x10^-300^* | *4.65* | *1.6x10^-116^* |
|  | *[5.94, 6.39]* |  | *[4.31, 4.99]* |  | *[6.55, 7.11]* |  | *[4.26, 5.05]* |  |
| COVID1 | -0.34 | **0.005** | -0.38 | 0.06 | -0.36 | **0.02** | -0.38 | 0.10 |
|  | [-0.57, -0.10] |  | [-0.78, 0.01] |  | [-0.65, -0.07] |  | [-0.83, 0.07] |  |
| COVID2 | -0.14 | 0.24 | 0.14 | 0.51 | -0.28 | 0.07 | 0.13 | 0.57 |
|  | [-0.39, 0.10] |  | [-0.26, 0.54] |  | [-0.58, 0.02] |  | [-0.32, 0.59] |  |
| COVID3 | -0.04 | 0.69 | 0.13 | 0.44 | -0.12 | 0.37 | 0.13 | 0.50 |
|  | [-0.25, 0.17] |  | [-0.20, 0.46] |  | [-0.38, 0.14] |  | [-0.25, 0.50] |  |
| ADHD | 5.14 | **6.6x10^-58^** | 3.76 | **5.7x10^-17^** | 6.21 | **2.5x10^-50^** | 3.76 | **6.1x10^-13^** |
|  | [4.51, 5.76] |  | [2.88, 4.63] |  | [5.39, 7.02] |  | [2.73, 4.78] |  |
| **COVID1*ADHD** | -2.10 | **1.5x10^-9^** | -2.16 | **4.2x10^-5^** | -2.11 | **2.1x10^-6^** | -2.16 | **0.0003** |
|  | [-2.78, -1.42] |  | [-3.19, -1.13] |  | [-2.98, -1.24] |  | [-3.35, -0.98] |  |
| **COVID2*ADHD** | -1.81 | **2.1x10^-7^** | -2.14 | **4.0x10^-5^** | -1.70 | **0.0002** | -2.14 | **0.0003** |
|  | [-2.50, -1.13] |  | [-3.16, -1.12] |  | [-2.59, -0.81] |  | [-3.31, -0.97] |  |
| **COVID3*ADHD** | -1.90 | **1.5x10^-10^** | -1.48 | **0.0005** | -2.15 | **3.4x10^-8^** | -1.48 | **0.003** |
|  | [-2.48, -1.32] |  | [-2.32, -0.64] |  | [-2.91, -1.38] |  | [-2.44, -0.52] |  |
| Sex |  |  |  |  |  |  | 2.18 | **3.4x10^-19^** |
|  |  |  |  |  |  |  | [1.70, 2.65] |  |
| COVID1*Sex |  |  |  |  |  |  | 0.02 | 0.93 |
|  |  |  |  |  |  |  | [-0.51, 0.55] |  |
| COVID2*Sex |  |  |  |  |  |  | -0.42 | 0.13 |
|  |  |  |  |  |  |  | [-0.96, 0.12] |  |
| COVID3*Sex |  |  |  |  |  |  | -0.25 | 0.28 |
|  |  |  |  |  |  |  | [-0.70, 0.20] |  |
| ADHD*Sex |  |  |  |  |  |  | 2.45 | **0.0002** |
|  |  |  |  |  |  |  | [1.17, 3.73] |  |
| COVID1*ADHD*Sex |  |  |  |  |  |  | 0.05 | 0.95 |
|  |  |  |  |  |  |  | [-1.40, 1.50] |  |
| COVID2*ADHD*Sex |  |  |  |  |  |  | 0.44 | 0.55 |
|  |  |  |  |  |  |  | [-1.00, 1.88] |  |
| COVID3*ADHD*Sex |  |  |  |  |  |  | -0.67 | 0.28 |
|  |  |  |  |  |  |  | [-1.87, 0.54] |  |
| **Random effects** | | | | | | | | |
| *Individual - Intercept* | 19.71 | **1.3x10^-214^** | 13.32 | **1.3x10^-63^** | 20.77 | **2.0x10^-147^** | 18.44 | **6.4x10^-209^** |
|  | [18.47, 20.94] |  | [11.77, 14.87] |  | [19.19, 22.34] |  | [17.27, 19.62] |  |
| *Timepoint - Intercept* | 14.41 | **<2.2x10^-300^** | 10.94 | **6.3x10^-220^** | 15.82 | **<2.2x10^-300^** | 14.39 | **<2.2x10^-300^** |
|  | [13.93, 14.89] |  | [10.27, 11.62] |  | [15.19, 16.45] |  | [13.90, 14.87] |  |
| **Model information** | | | | | | | | |
| N | 9886 | | 2954 | | 6932 | | 9886 | |
| logLik | -29757.54 | |  | |  | | -29670.11 | |
| AIC | 59535.09 | |  | |  | | 59376.22 | |
| BIC | 59607.07 | |  | |  | | 59505.80 | |

**Supplementary Table 4** - *Multilevel model of depression and ASD. Beta coefficients [95% CIs] are presented for each variable in the four models. Positive beta coefficients indicate worse depressive symptoms.*

| **Depression** | **Model 1**  Total | | **Model 2**  Male strata | | **Model 3**  Female strata | | **Model 4**  Total, sex-interaction | |
| --- | --- | --- | --- | --- | --- | --- | --- | --- |
|  | Beta coefficient  [95% CIs] | P-value | Beta coefficient  [95% CIs] | P-value | Beta coefficient  [95% CIs] | P-value | Beta coefficient  [95% CIs] | P-value |
| **Fixed effects** | | | | | | | | |
| *Intercept* | *6.63* | *<2.2x10^-300^* | *5.11* | *7.7x10^-211^* | *7.32* | *<2.2x10^-300^* | *5.11* | *5.5x10^-155^* |
|  | *[6.42, 6.85]* |  | *[4.78, 5.43]* |  | *[7.06, 7.59]* |  | *[4.73, 5.48]* |  |
| COVID1 | -0.52 | **5.2x10^-6^** | -0.68 | **0.0003** | -0.50 | **0.00041** | -0.68 | **0.002** |
|  | [-0.75, -0.30] |  | [-1.06, -0.31] |  | [-0.78, -0.22] |  | [-1.11, -0.26] |  |
| COVID2 | -0.30 | **0.01** | -0.14 | 0.48 | -0.40 | **0.006** | -0.14 | 0.53 |
|  | [-0.53, -0.07] |  | [-0.51, 0.24] |  | [-0.69, -0.11] |  | [-0.57, 0.29] |  |
| COVID3 | -0.22 | **0.03** | -0.07 | 0.67 | -0.29 | **0.02** | -0.07 | 0.71 |
|  | [-0.42, -0.03] |  | [-0.38, 0.24] |  | [-0.54, -0.05] |  | [-0.42, 0.28] |  |
| ASD | 7.21 | **2.6x10^-26^** | 4.19 | **2.9x10^-5^** | 8.74 | **3.7x10^-24^** | 4.18 | **0.0004** |
|  | [5.88, 8.55] |  | [2.22, 6.15] |  | [7.05, 10.44] |  | [1.89, 6.48] |  |
| **COVID1*ASD** | -3.09 | **1.6x10^-5^** | -0.85 | 0.44 | -4.10 | **6.6x10^-6^** | -0.84 | 0.51 |
|  | [-4.49, -1.69] |  | [-3.01, 1.31] |  | [-5.88, -2.32] |  | [-3.31, 1.63] |  |
| **COVID2*ASD** | -2.83 | **4.9x10^-5^** | -1.86 | 0.08 | -3.27 | **0.0002** | -1.87 | 0.13 |
|  | [-4.20, -1.47] |  | [-3.98, 0.26] |  | [-5.01, -1.54] |  | [-4.29, 0.55] |  |
| **COVID3*ASD** | -2.34 | **0.0002** | -0.88 | 0.35 | -3.05 | **0.0001** | -0.88 | 0.42 |
|  | [-3.56, -1.12] |  | [-2.74, 0.98] |  | [-4.61, -1.49] |  | [-3.00, 1.24] |  |
| Sex |  |  |  |  |  |  | 2.22 | **1.3x10^-21^** |
|  |  |  |  |  |  |  | [1.76, 2.67] |  |
| COVID1*Sex |  |  |  |  |  |  | 0.18 | 0.48 |
|  |  |  |  |  |  |  | [-0.32, 0.68] |  |
| COVID2*Sex |  |  |  |  |  |  | -0.26 | 0.31 |
|  |  |  |  |  |  |  | [-0.77, 0.25] |  |
| COVID3*Sex |  |  |  |  |  |  | -0.23 | 0.30 |
|  |  |  |  |  |  |  | [-0.65, 0.20] |  |
| ASD*Sex |  |  |  |  |  |  | 4.56 | **0.001** |
|  |  |  |  |  |  |  | [1.77, 7.36] |  |
| COVID1*ASD*Sex |  |  |  |  |  |  | -3.26 | **0.03** |
|  |  |  |  |  |  |  | [-6.26, -0.26] |  |
| COVID2*ASD*Sex |  |  |  |  |  |  | -1.40 | 0.35 |
|  |  |  |  |  |  |  | [-4.33, 1.53] |  |
| COVID3*ASD*Sex |  |  |  |  |  |  | -2.18 | 0.10 |
|  |  |  |  |  |  |  | [-4.77, 0.42] |  |
| **Random effects** | | | | | | | | |
| *Individual - Intercept* | 20.56 | **8.6x10^-218^** | 13.78 | **3.4x10^-64^** | 22.09 | **6.4x10^-151^** | 19.49 | **2.5x10^-213^** |
|  | [19.28, 21.83] |  | [12.18, 15.37] |  | [20.43, 23.74] |  | [18.27, 20.72] |  |
| *Timepoint - Intercept* | 14.48 | **<2.2x10^-300^** | 11.08 | **6.3x10^-220^** | 15.86 | **<2.2x10^-300^** | 14.46 | **<2.2x10^-300^** |
|  | [14.00, 14.97] |  | [10.39, 11.76] |  | [15.23, 16.50] |  | [13.97, 14.94] |  |
| **Model information** | | | | | | | | |
| N | 9886 | | 2954 | | 6932 | | 9886 | |
| logLik | -29827.45 | |  | |  | | -29755.57 | |
| AIC | 59674.89 | |  | |  | | 59547.14 | |
| BIC | 59746.88 | |  | |  | | 59676.72 | |

**Supplementary Table 5** - *Multilevel model of mental wellbeing and ADHD. Beta coefficients [95% CIs] are presented for each variable in the four models. Negative beta coefficients indicate worse mental wellbeing.*

| **Mental Wellbeing** | **Model 1**  Total | | **Model 2**  Male strata | | **Model 3**  Female strata | | **Model 4**  Total, sex-interaction | |
| --- | --- | --- | --- | --- | --- | --- | --- | --- |
|  | Beta coefficient  [95% CIs] | P-value | Beta coefficient  [95% CIs] | P-value | Beta coefficient  [95% CIs] | P-value | Beta coefficient  [95% CIs] | P-value |
| **Fixed effects** | | | | | | | | |
| *Intercept* | *49.56* | *<2.2x10^-300^* | *50.40* | *<2.2x10^-300^* | *49.20* | *<2.2x10^-300^* | *50.39* | *<2.2x10^-300^* |
|  | *[49.19, 49.94]* |  | *[49.70, 51.09]* |  | *[48.75, 49.64]* |  | *[49.70, 51.09]* |  |
| COVID1 | -4.97 | **1.6x10^-130^** | -4.28 | **2.3x10^-27^** | -5.22 | **6.9x10^-106^** | -4.28 | **1.4x10^-27^** |
|  | [-5.37, -4.57] |  | [-5.06, -3.51] |  | [-5.68, -4.75] |  | [-5.05, -3.51] |  |
| COVID2 | -4.68 | **6.8x10^-111^** | -4.70 | **1.4x10^-31^** | -4.67 | **2.7x10^-81^** | -4.70 | **8.2x10^-32^** |
|  | [-5.09, -4.27] |  | [-5.49, -3.91] |  | [-5.15, -4.19] |  | [-5.48, -3.91] |  |
| COVID3 | -4.15 | **3.2x10^-112^** | -3.62 | **5.2x10^-26^** | -4.37 | **1.3x10^-89^** | -3.62 | **3.2x10^-26^** |
|  | [-4.51, -3.78] |  | [-4.29, -2.95] |  | [-4.80, -3.95] |  | [-4.29, -2.95] |  |
| ADHD | -5.12 | **5.8x10^-21^** | -4.22 | **3.5x10^-6^** | -5.80 | **1.1x10^-17^** | -4.22 | **3.7x10^-6^** |
|  | [-6.19, -4.05] |  | [-6.01, -2.44] |  | [-7.13, -4.47] |  | [-6.00, -2.43] |  |
| **COVID1*ADHD** | 1.30 | **0.03** | 0.78 | 0.45 | 1.50 | **0.04** | 0.78 | 0.45 |
|  | [0.15, 2.46] |  | [-1.25, 2.81] |  | [0.09, 2.90] |  | [-1.25, 2.80] |  |
| **COVID2*ADHD** | 1.27 | **0.03** | 1.21 | 0.24 | 1.32 | 0.07 | 1.21 | 0.24 |
|  | [0.11, 2.44] |  | [-0.80, 3.22] |  | [-0.11, 2.76] |  | [-0.79, 3.21] |  |
| **COVID3*ADHD** | 0.20 | 0.70 | 0.27 | 0.76 | 0.07 | 0.92 | 0.27 | 0.76 |
|  | [-0.82, 1.23] |  | [-1.46, 2.00] |  | [-1.21, 1.34] |  | [-1.46, 1.99] |  |
| Sex |  |  |  |  |  |  | -1.20 | **0.005** |
|  |  |  |  |  |  |  | [-2.02, -0.37] |  |
| COVID1*Sex |  |  |  |  |  |  | -0.93 | **0.04** |
|  |  |  |  |  |  |  | [-1.84, -0.03] |  |
| COVID2*Sex |  |  |  |  |  |  | 0.03 | 0.95 |
|  |  |  |  |  |  |  | [-0.89, 0.95] |  |
| COVID3*Sex |  |  |  |  |  |  | -0.75 | 0.06 |
|  |  |  |  |  |  |  | [-1.55, 0.04] |  |
| ADHD*Sex |  |  |  |  |  |  | -1.58 | 0.16 |
|  |  |  |  |  |  |  | [-3.81, 0.64] |  |
| COVID1*ADHD*Sex |  |  |  |  |  |  | 0.72 | 0.57 |
|  |  |  |  |  |  |  | [-1.75, 3.18] |  |
| COVID2*ADHD*Sex |  |  |  |  |  |  | 0.11 | 0.93 |
|  |  |  |  |  |  |  | [-2.35, 2.57] |  |
| COVID3*ADHD*Sex |  |  |  |  |  |  | -0.20 | 0.85 |
|  |  |  |  |  |  |  | [-2.34, 1.94] |  |
| **Random effects** | | | | | | | | |
| *Individual - Intercept* | 43.98 | **4.3x10^-195^** | 42.49 | **1.6x10^-58^** | 43.57 | **5.0x10^-137^** | 43.25 | **1.0x10^-193^** |
|  | [41.08, 46.87] |  | [37.33, 47.66] |  | [40.14, 47.00] |  | [40.40, 46.11] |  |
| *Timepoint - Intercept* | 35.46 | **<2.2x10^-300^** | 35.74 | **6.8x10^-191^** | 35.25 | **<2.2x10^-300^** | 35.39 | **<2.2x10^-300^** |
|  | [34.20, 36.71] |  | [33.36, 38.11] |  | [33.78, 36.73] |  | [34.13, 36.64] |  |
| **Model information** | | | | | | | | |
| N | 9143 | | 2678 | | 6465 | | 9143 | |
| logLik | -31590.39 | |  | |  | | -31564.42 | |
| AIC | 63200.77 | |  | |  | | 63164.83 | |
| BIC | 63271.98 | |  | |  | | 63293.00 | |

**Supplementary Table 6** - *Multilevel model of mental wellbeing and ASD. Beta coefficients [95% CIs] are presented for each variable in the four models. Negative beta coefficients indicate worse mental wellbeing.*

| **Mental Wellbeing** | **Model 1**  Total | | **Model 2**  Male strata | | **Model 3**  Female strata | | **Model 4**  Total, sex-interaction | |
| --- | --- | --- | --- | --- | --- | --- | --- | --- |
|  | Beta coefficient  [95% CIs] | P-value | Beta coefficient  [95% CIs] | P-value | Beta coefficient  [95% CIs] | P-value | Beta coefficient  [95% CIs] | P-value |
| **Fixed effects** | | | | | | | | |
| *Intercept* | *49.18* | *<2.2x10^-300^* | *49.95* | *<2.2x10^-300^* | *48.84* | *<2.2x10^-300^* | *49.95* | *<2.2x10^-300^* |
|  | *[48.83, 49.54]* |  | *[49.30, 50.60]* |  | *[48.41, 49.26]* |  | *[49.30, 50.60]* |  |
| COVID1 | -4.86 | **6.8x10^-138^** | -4.11 | **1.4x10^-28^** | -5.14 | **2.5x10^-112^** | -4.11 | **8.6x10^-29^** |
|  | [-5.24, -4.48] |  | [-4.84, -3.38] |  | [-5.58, -4.69] |  | [-4.83, -3.39] |  |
| COVID2 | -4.57 | **4.4x10^-117^** | -4.52 | **2.6x10^-33^** | -4.59 | **8.9x10^-86^** | -4.52 | **1.5x10^-33^** |
|  | [-4.96, -4.18] |  | [-5.25, -3.78] |  | [-5.04, -4.13] |  | [-5.25, -3.78] |  |
| COVID3 | -4.17 | **1.4x10^-125^** | -3.59 | **3.6x10^-29^** | -4.42 | **3.7x10^-100^** | -3.59 | **2.1x10^-29^** |
|  | [-4.51, -3.82] |  | [-4.22, -2.97] |  | [-4.83, -4.01] |  | [-4.22, -2.97] |  |
| ASD | -9.96 | **5.2x10^-19^** | -7.19 | **0.0004** | -11.18 | **4.8x10^-17^** | -7.18 | **0.0004** |
|  | [-12.15, -7.77] |  | [-11.17, -3.20] |  | [-13.79, -8.57] |  | [-11.17, -3.19] |  |
| **COVID1*ASD** | 1.82 | 0.13 | -1.12 | 0.61 | 3.02 | **0.04** | -1.13 | 0.60 |
|  | [-0.55, 4.20] |  | [-5.40, 3.16] |  | [0.17, 5.88] |  | [-5.39, 3.14] |  |
| **COVID2*ASD** | 1.80 | 0.13 | 0.63 | 0.77 | 2.26 | 0.11 | 0.63 | 0.76 |
|  | [-0.52, 4.12] |  | [-3.52, 4.79] |  | [-0.53, 5.06] |  | [-3.50, 4.77] |  |
| **COVID3*ASD** | 1.22 | 0.26 | 0.43 | 0.83 | 1.45 | 0.25 | 0.42 | 0.83 |
|  | [-0.88, 3.32] |  | [-3.44, 4.29] |  | [-1.05, 3.95] |  | [-3.42, 4.27] |  |
| Sex |  |  |  |  |  |  | -1.11 | **0.005** |
|  |  |  |  |  |  |  | [-1.89, -0.33] |  |
| COVID1*Sex |  |  |  |  |  |  | -1.03 | **0.018** |
|  |  |  |  |  |  |  | [-1.88, -0.18] |  |
| COVID2*Sex |  |  |  |  |  |  | -0.07 | 0.88 |
|  |  |  |  |  |  |  | [-0.93, 0.80] |  |
| COVID3*Sex |  |  |  |  |  |  | -0.83 | **0.03** |
|  |  |  |  |  |  |  | [-1.58, -0.08] |  |
| ASD*Sex |  |  |  |  |  |  | -4.00 | 0.10 |
|  |  |  |  |  |  |  | [-8.77, 0.77] |  |
| COVID1*ASD*Sex |  |  |  |  |  |  | 4.15 | 0.11 |
|  |  |  |  |  |  |  | [-0.98, 9.28] |  |
| COVID2*ASD*Sex |  |  |  |  |  |  | 1.63 | 0.52 |
|  |  |  |  |  |  |  | [-3.37, 6.62] |  |
| COVID3*ASD*Sex |  |  |  |  |  |  | 1.03 | 0.66 |
|  |  |  |  |  |  |  | [-3.56, 5.62] |  |
| **Random effects** | | | | | | | | |
| *Individual - Intercept* | 44.30 | **1.1x10^-195^** | 43.02 | **7.1x10^-59^** | 44.02 | **1.3x10^-137^** | 43.72 | **1.2x10^-194^** |
|  | [41.39, 47.21] |  | [37.81, 48.23] |  | [40.56, 47.47] |  | [40.84, 46.60] |  |
| *Timepoint - Intercept* | 35.48 | **<2.2x10^-300^** | 35.73 | **7.1x10^-191^** | 35.27 | **<2.2x10^-300^** | 35.40 | **<2.2x10^-300^** |
|  | [34.22, 36.73] |  | [33.35, 38.11] |  | [33.79, 36.75] |  | [34.14, 36.65] |  |
| **Model information** | | | | | | | | |
| N | 9143 | | 2678 | | 6465 | | 9143 | |
| logLik | -31601.13 | |  | |  | | -31578.09 | |
| AIC | 63222.26 | |  | |  | | 63192.18 | |
| BIC | 63293.47 | |  | |  | | 63320.35 | |

**Supplementary Table 7** - *Cohort information stratified using different classifications of ADHD and ASD.* P-values from Chi-squared tests between groups are presented.

|  | **ADHD (SDQ ≥ 5)** | | | **ADHD (SDQ ≥ 7)** | | | **ASD (ASQ ≥ 27)** | | |
| --- | --- | --- | --- | --- | --- | --- | --- | --- | --- |
|  | **Yes** | **No** | **P-value** | **Yes** | **No** | **P-value** | **Yes** | **No** | **P-value** |
| **N** | 747 (24) | 2311 (75) | - | 200 (7) | 2858 (93) | - | 336 (11) | 2722 (89) | - |
| **Sex** |  |  |  |  |  |  |  |  |  |
| Females (%) | 468 (63) | 1,635 (71) | 3.3x10^-5^ | 118 (59) | 1,985(69) | 0.002 | 197 (59) | 1,906 (70) | 2.1x10^-5^ |
| **ADHD symptoms** |  |  |  |  |  |  |  |  |  |
| Mean SDQ score (SD) | 5.94 (1.15) | 2.10 (1.35) | - | 7.58 (0.82) | 2.72 (1.77) | - | 4.38 (2.31) | 2.87 (2.02) | - |
| ADHD based on SDQ ≥ 5 (%) | 747 (100) | 0 (0) | - | 200(100) | 547 (19) | - | 162 (48)^^ | 585 (21) | 5.6x10^-27^ |
| ADHD based on SDQ ≥ 7 (%) | 200 (27) | 0 (0) | - | 200 (100) | 0 (0) | - | 62 (18) | 138 (5) | 8.0x10^-21^ |
| **ASD symptoms** |  |  |  |  |  |  |  |  |  |
| Mean AQ score (SD) | 19.49 (7.75) | 14.86 (6.62) |  | 21.90 (7.57) | 15.58 (6.98) | - | 29.88 (3.95) | 14.28 (5.42) | - |
| ASD based on AQ ≥ 27 (%) | 162 (22) | 174 (8) | 5.6x10^-27^ | 62 (31) | 274 (10) | 8.0x10^-21^ | 336 (100) | 0 (0) | - |
| **Events during pandemic** |  |  |  |  |  |  |  |  |  |
| Financial difficulties (%) | 235 (31) | 529 (23) | 2.6x10^-6^ | 71 (36) | 693 (24) | 0.0004 | 102 (30) | 662 (24) | 0.02 |
| Lost job (%) | 71 (10) | 168 (7) | 0.05 | 20 (10) | 219 (8) | 0.23 | 27 (8) | 212 (8) | 0.87 |
| Death of someone close (%) | 141 (19) | 392 (17) | 0.23 | 45 (22) | 488 (17) | 0.05 | 57 (17) | 476 (17) | 0.81 |
| Illness/injury of someone  close (%) | 130 (17) | 384 (17) | 0.62 | 39 (20) | 475 (17) | 0.29 | 59 (18) | 455 (17) | 0.70 |
| **Anxiety (GAD7)** |  |  |  |  |  |  |  |  |  |
| Pre-pandemic (%) | 101/463 (22) | 154/1,535 (10) | 2.8x10^-11^ | 31/112 (28) | 224/1,886 (12) | 1.1x10^-6^ | 58/204 (28) | 197/1,794 (12) | 1.5x10^-12^ |
| COVID1 (%) | 165/484 (34) | 326/1,564 (21) | 2.4x10^-9^ | 49/125 (39) | 442/1,923 (23) | 3.9x10^-5^ | 93/231 (40) | 398/1,817 (22) | 7.5x10^-10^ |
| COVID2 (%) | 168/452 (37) | 269/1,445 (19) | 2.9x10^-16^ | 50/121 (41) | 387/1,776 (22) | 7.9x10^-7^ | 95/212 (45) | 342/1,685 (20) | 1.4x10^-15^ |
| COVID3 (%) | 282/738 (38) | 538/2,290 (23) | 5.1x10^-15^ | 85/200 (43) | 735/2,828 (26) | 3.8x10^-7^ | 136/334 (41) | 684/2,694 (25) | 2.7x10^-9^ |
| **Depression (SMFQ)** |  |  |  |  |  |  |  |  |  |
| Pre-pandemic (%) | 266/732 (36) | 349/2,241 (16) | 2.1x10^-33^ | 94/194 (48) | 521/2,779 (19) | 5.3x10^-23^ | 160/329 (49) | 455/2,644 (17) | 4.5x10^-40^ |
| COVID1 (%) | 114/474 (24) | 185/1,550 (12) | 7.8x10^-11^ | 39/123 (32) | 260/1,901 (14) | 4.7x10^-8^ | 66/228 (29) | 233/1,796 (13) | 1.5x10^-10^ |
| COVID2 (%) | 149/445 (33) | 191/1,447 (13) | 1.9x10^-22^ | 43/120 (36) | 297/1,772 (17) | 1.4x10^-7^ | 85/213 (40) | 255/1,679 (15) | 8.6x10^-19^ |
| COVID3 (%) | 213/734 (29) | 345/2,263 (15) | 8.1x10^-17^ | 75/199 (38) | 483/2,798 (17) | 8.5x10^-13^ | 106/331 (32) | 452/2,666 (17) | 3.1x10^-11^ |
| **Poor mental wellbeing (WEMWBS)** |  |  |  |  |  |  |  |  |  |
| Pre-pandemic (%) | 168/518 (32) | 245/1,674 (15) | 1.4x10^-19^ | 52/134 (39) | 361/2,058 (18) | 1.1x10^-9^ | 103/245 (42) | 310/1,947 (16) | 6.6x10^-23^ |
| COVID1 (%) | 203/479 (42) | 425/1,556 (27) | 4.3x10^-10^ | 62/121 (51) | 566/1,914 (30) | 5.6x10^-7^ | 124/229 (54) | 504/1,806 (28) | 5.6x10^-16^ |
| COVID2 (%) | 214/449 (48) | 388/1,453 (27) | 7.1x10^-17^ | 60/121 (50) | 542/1,781 (30) | 1.2x10^-5^ | 127/213 (60) | 475/1,689 (28) | 1.2x10^-20^ |
| COVID3 (%) | 322/736 (44) | 606/2,278 (27) | 1.9x10^-18^ | 103/198 (52) | 825/2,816 (29) | 2.2x10^-11^ | 177/331 (53) | 751/2,683 (28) | 2.6x10^-21^ |

|  | | **GAD7** | | | | **SMFQ** | | | | **WEMWBS** | | | |
| --- | --- | --- | --- | --- | --- | --- | --- | --- | --- | --- | --- | --- | --- |
|  |  | **Pre-COVID** | **COVID**  **1** | **COVID**  **2** | **COVID**  **3** | **Pre-COVID** | **COVID**  **1** | **COVID**  **2** | **COVID**  **3** | **Pre-COVID** | **COVID**  **1** | **COVID**  **2** | **COVID**  **3** |
| **GAD7** | **Pre-COVID** | 1 | 0.41 | 0.40 | 0.47 | 0.49 | 0.36 | 0.36 | 0.43 | -0.38 | -0.32 | -0.33 | -0.36 |
|  | **COVID1** | 0.41 | 1 | 0.74 | 0.64 | 0.48 | 0.76 | 0.62 | 0.56 | -0.32 | -0.59 | -0.52 | -0.49 |
|  | **COVID2** | 0.40 | 0.74 | 1 | 0.66 | 0.50 | 0.64 | 0.78 | 0.58 | -0.35 | -0.52 | -0.64 | -0.50 |
|  | **COVID3** | 0.47 | 0.64 | 0.66 | 1 | 0.48 | 0.54 | 0.56 | 0.76 | -0.33 | -0.44 | -0.49 | -0.66 |
| **SMFQ** | **Pre-COVID** | 0.49 | 0.48 | 0.50 | 0.48 | 1 | 0.54 | 0.54 | 0.53 | -0.46 | -0.47 | -0.48 | -0.45 |
|  | **COVID1** | 0.36 | 0.76 | 0.64 | 0.54 | 0.54 | 1 | 0.72 | 0.62 | -0.34 | -0.66 | -0.58 | -0.51 |
|  | **COVID2** | 0.36 | 0.62 | 0.78 | 0.56 | 0.54 | 0.72 | 1 | 0.66 | -0.37 | -0.56 | -0.72 | -0.56 |
|  | **COVID3** | 0.43 | 0.56 | 0.58 | 0.76 | 0.53 | 0.62 | 0.66 | 1 | -0.36 | -0.48 | -0.54 | -0.70 |
| **WEMWBS** | **Pre-COVID** | -0.38 | -0.32 | -0.35 | -0.33 | -0.46 | -0.34 | -0.37 | -0.36 | 1 | 0.46 | 0.48 | 0.45 |
|  | **COVID1** | -0.32 | -0.59 | -0.52 | -0.44 | -0.47 | -0.66 | -0.56 | -0.48 | 0.46 | 1 | 0.72 | 0.61 |
|  | **COVID2** | -0.33 | -0.52 | -0.64 | -0.49 | -0.48 | -0.58 | -0.72 | -0.54 | 0.48 | 0.72 | 1 | 0.67 |
|  | **COVID3** | -0.36 | -0.49 | -0.50 | -0.66 | -0.45 | -0.51 | -0.56 | -0.70 | 0.45 | 0.61 | 0.67 | 1 |

**Supplementary Table 8** - *Correlation between each mental health questionnaire at each timepoint. Spearman’s r coefficient presented. Since group sizes are different at each timepoint, pairwise exclusion was performed for each correlation*
